# How immunity shapes the long-term dynamics of seasonal influenza

**DOI:** 10.1101/2023.09.08.23295244

**Authors:** Oliver Eales, Freya Shearer, James McCaw

## Abstract

Since its emergence in 1968, influenza A H3N2 has caused yearly epidemics in temperate regions. While infection confers immunity against antigenically similar strains, new antigenically distinct strains that evade existing immunity regularly emerge (‘antigenic drift’). Immunity at the individual level is complex, depending on an individual’s lifetime infection history. An individual’s first infection with influenza typically elicits the greatest response with subsequent infections eliciting progressively reduced responses (‘antigenic seniority’). The combined effect of individual-level immune responses and antigenic drift on the epidemiological dynamics of influenza are not well understood. Here we develop an integrated modelling framework of influenza transmission, immunity, and antigenic drift to show how individual-level exposure, and the build-up of population level immunity, shape the long-term epidemiological dynamics of H3N2. Including antigenic seniority in the model, we observe that following an initial decline after the pandemic year, the average annual attack rate increases over the next 80 years, before reaching an equilibrium, with greater increases in older age-groups. Our analyses suggest that the average attack rate of H3N2 is still in a growth phase. Further increases, particularly in the elderly, may be expected in coming decades, driving an increase in healthcare demand due to H3N2 infections.

We anticipate our findings and methodological developments will be applicable to other antigenically variable pathogens. This includes the recent pandemic pathogens influenza A H1N1pdm09, circulating since 2009, and SARS-CoV-2, circulating since 2019. Our findings highlight that following the short-term reduction in attack rates after a pandemic, if there is any degree of antigenic seniority then a resurgence in attack rates should be expected over the longer-term. Designing and implementing studies to assess the dynamics of immunity for H1N1pdm09, SARS-CoV-2, and other antigenically variable pathogens may help anticipate any long-term rises in infection and health burden.

## Introduction

Influenza A H3N2 has circulated in the human population since its emergence in 1968 (the ‘Hong Kong’ pandemic), causing substantial mortality and morbidity [1]. Infection with a strain of influenza typically causes an immune response, in which antibodies are produced that can recognise and bind to antigenic sites on the virus’ surface proteins. These antibodies protect against future infection with antigenically similar strains of influenza. However, due to the accumulation of mutations, new antigenically distinct strains regularly emerge that are able to evade existing immunity (‘antigenic drift’) [2].

In temperate regions, where influenza transmission is highly seasonal, there are yearly winter epidemics, but during the summer (when there is little transmission) strains regularly become extinct [3]. In contrast, the tropics experience a more continuous transmission regime [4], with more consistent levels of infection throughout the year. As a result, epidemics of new antigenic variants in temperate regions are predominantly seeded by tropical regions [5], particularly East and South-East Asia which has a major role in the production and dissemination of new antigenic variants [6].

Antigenic cartography has been used extensively in assessing the antigenic evolution of influenza H3N2 [1,7]. The antigenic similarity between two strains can be assessed using the haemagglutinin inhibition (HI) assay; this measures the ability of antisera raised against one strain of influenza to prevent agglutination of red blood cells by another strain of influenza (proportional to the antibody concentration against the strain). Through pairwise comparisons of the HI assay across many strains the antigenic distance between strains can be inferred and represented on a two-dimensional ‘antigenic map’ [1,7].

At the individual level, antibody concentration (against a specific strain) is related to the individual’s level of protection against infection (when exposed to that strain) [8] (Fig.1C). An individual’s antibody dynamics in response to exposure are complex, depending on their entire infection history. Following infection, antibody responses generated are greatest against the strain of infection, while they are reduced against (antigenically) distant strains. A broadly cross-protective short-term antibody response fades to leave a more specific long-term response [9] (Fig.1A). An individual’s first infection with influenza typically elicits the greatest antibody response and subsequent infections elicit progressively reduced antibody responses [10] (Fig.1B). This progressive reduction is known as ‘antigenic seniority’.

**Figure 1.**
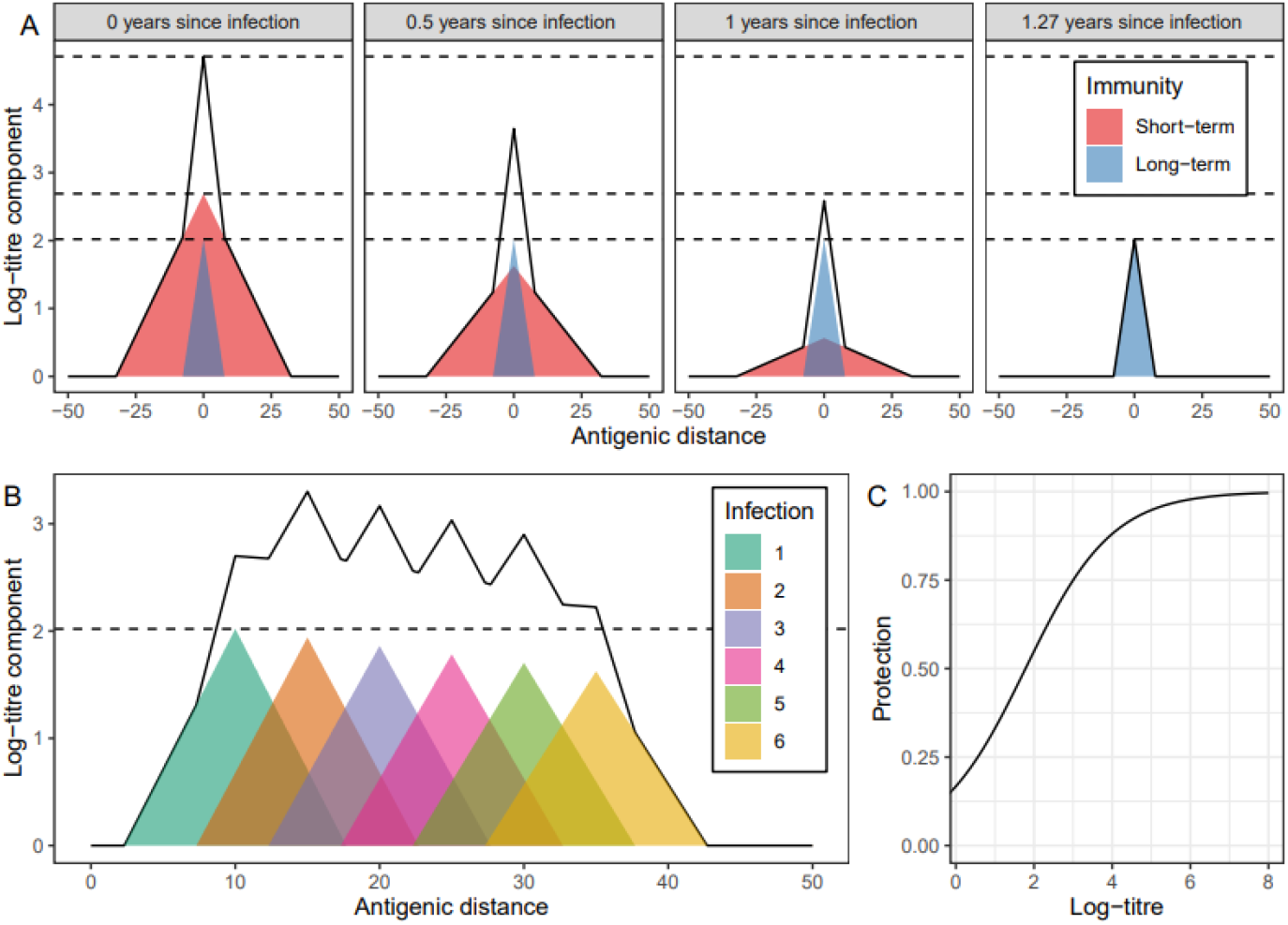
Relationship between infection and immunity. **(A)** Short- (red) and long-term (blue) components of the antibody response (measured by log-titre of a haemagglutinin inhibition test), and the total response (solid black line), due to an infection with a strain of influenza. Antibody response is greatest against the strain of infection (antigenic distance = 0) and decreases linearly with antigenic distance against other strains. The antigenic distance between two strains quantifies their dissimilarity, with one unit of antigenic distance corresponding to a two-fold dilution in hemagglutinin inhibition (HI) titre. This HI titre measures the ability of antisera raised against one strain of influenza to prevent agglutination of red blood cells by another strain of influenza.. The short-term response decreases linearly with time, reaching zero 1.27 years after infection. **(B)** Long-term components of the antibody response (coloured), and total long-term antibody response (solid black line), due to infection with multiple strains of influenza (evenly spaced in one-dimension of antigenic space). The first infection has the greatest antibody response (dashed black line shows the maximum value [against strain of first infection]), subsequent infections have suppressed antibody responses, decreasing linearly with the number of previous infections. **(C)** Relationship between the log-titre against a specific strain of influenza, and the protection against infection with that strain. Note that due to our parameterisation (see Methods) the protection against infection does not go to zero for a log-titre of zero. This is not a problem as only the relative values of protection against infection (by log-titre) are of concern — the non-zero value simply decreases the transmission rate which we have nevertheless adjusted to fit observed data.

Epidemic models can be used to explore the dynamics of multi-strain pathogens such as influenza. In these multi-strain models a ‘strain space’ is defined; this space encompasses all possible strains that can circulate and describes the degree of cross-immunity between any pair of strains [11–13]. Such models have previously been used to explore the evolutionary dynamics of H3N2, incorporating mutation explicitly into the transmission process. However, these models have focused on how mechanisms of immunity can give rise to the evolutionary patterns of H3N2; less consideration has been given to how evolutionary patterns at the global scale and immune mechanisms at the individual-level influence population-level epidemiological dynamics. Furthermore, the role of antigenic seniority, the magnitude of which has only recently been estimated [9,14], has not yet been investigated.

Here we develop an integrated modelling framework of influenza transmission, immunity, and antigenic evolution to study the long-term epidemiological dynamics of influenza and explore their dependence on different hypothesised and studied mechanisms of immunity [9,14], including antigenic seniority. We simulate the yearly epidemic dynamics of influenza transmission within a temperate region over a 160-year period. The strains present in the model are defined over a two-dimensional strain space [11,15], analogous to two-dimensional antigenic maps and tuned to accurately resemble the observed H3N2 maps. The degree of cross-immunity between strains is determined by their antigenic distance and a function linking their antigenic distance to immunity, based on previously published work [8,9] (Fig.1). Finally, antigenic evolution is introduced as a global process, enabling us to explore how evolutionary dynamics, controlled by tropical regions, influence the epidemiological dynamics of influenza in temperate regions.

### Simulating the long-term dynamics of influenza

We simulated the antigenic evolution of influenza H3N2 using a statistical model (see Methods) that describes the within- and between-year variation in antigenic diversity. The model’s parameters were fitted to available data from antigenic maps from 1968 to 2011 (Sup Fig.1) [1]. Our model allows us to simulate the strains of influenza present in a given epidemic year (N=20 per year) by their location on a two-dimensional antigenic map (Fig.2). We used the model to simulate many different evolutionary trajectories (Sup Fig.2) that resemble the punctuated nature of antigenic drift observed in data [7].

**Figure 2.**
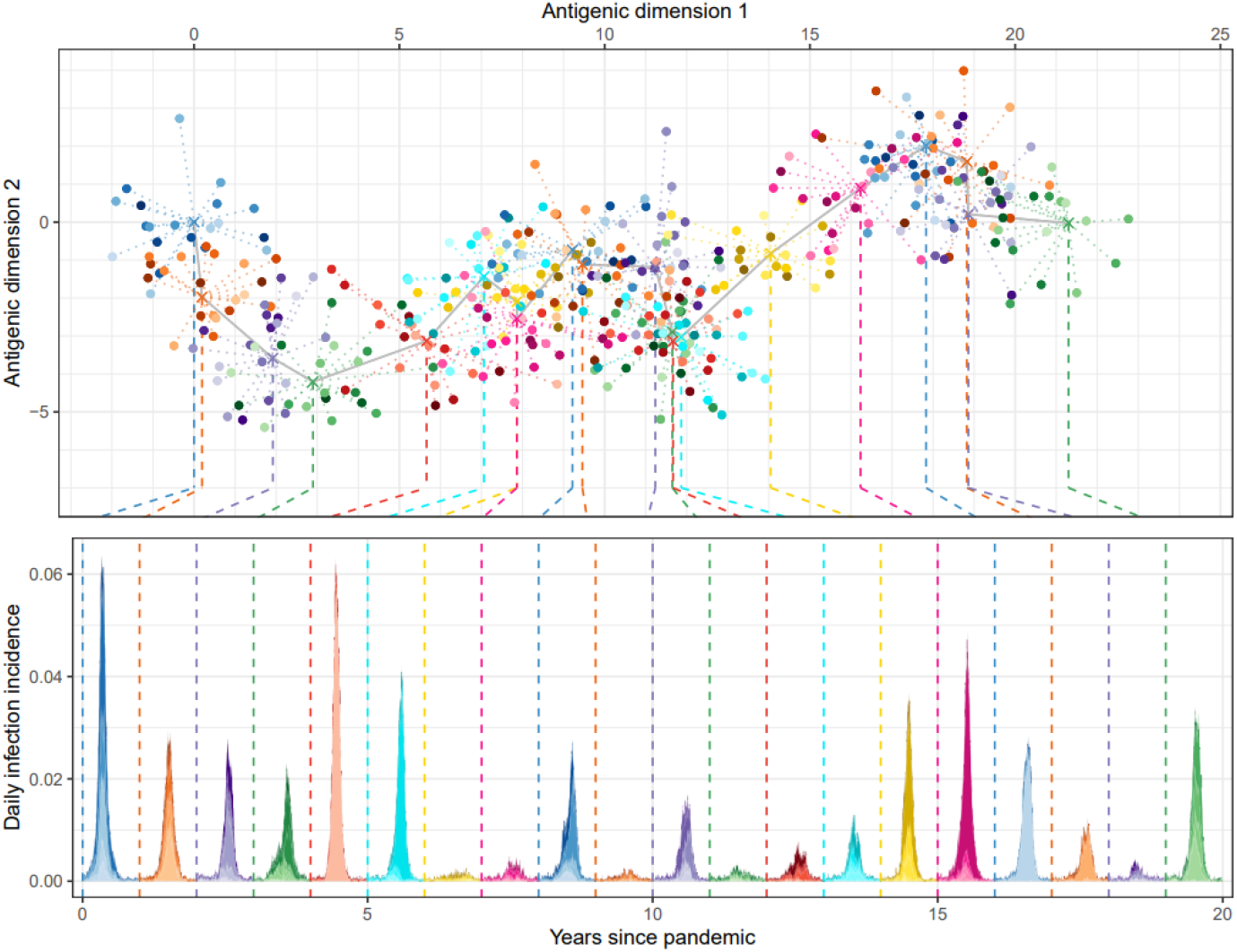
Simulated antigenic drift and epidemic dynamics of influenza A H3N2. (Top) Simulated antigenic diversity of influenza strains for 20 years (colours show separate years, with shade of colour highlighting different strains [20 per year]). Strains are located on a two-dimensional antigenic map. The antigenic distance between two strains quantifies their dissimilarity, with one unit of antigenic distance corresponding to a two-fold dilution in hemagglutinin inhibition titre. Strains in a single year (points) are normally distributed around mean antigenic coordinates (crosses, dotted lines connecting to points). The mean antigenic coordinates change each year (grey line connecting adjacent years), always progressing along antigenic dimension 1 (left to right). The mean antigenic coordinates in the first year were (0, 0). (Bottom) The resulting epidemic dynamics due to the 20 strains present in each year (dashed line connects strains in top panel to the beginning of the year they were present in in the bottom panel). The daily infection incidence for each strain (different shades of single colour) and the overall daily infection incidence (total shaded area) are shown for each discrete year (different colours between adjacent years).

Strains in the model — which incorporates seasonality and realistic contact patterns between age-groups (see Methods) — were introduced into a multi-strain agent-based epidemic model at a constant rate. The infection history of each individual in the model was tracked and used to calculate their probability of infection given contact with an individual infected with a specific strain (Fig.1, Sup Fig.3). Parameter values were selected from previous studies where available or chosen to best match observed data (See Methods and Sup Tab. 3). The resulting dynamics resembled the recurrent epidemics of influenza transmission observed in temperate regions (Fig.2). Between years there was substantial variation in the epidemic size, but limited variation in the timing of the epidemic peak (due to seasonality). Low levels of infection were observed in years with limited antigenic drift and high levels of infection were observed following years with substantial antigenic transitions. Following the buildup of population immunity in the years post emergence, epidemics showed little strain diversity during years with substantial antigenic diversity, with the most antigenically distinct strain dominating the epidemic dynamics (see year 4 in Fig.2).

There was substantial variation in the year-to-year annual attack rate (total proportion of population infected) between simulations (Sup Fig.4), but good agreement in the overall distribution of yearly attack rates (Sup Fig.4C). For this reason we considered the dynamics of the average annual attack rate across simulations.

### Long-term changes in influenza attack rates

The average annual attack rate changed substantially over 160 years since introduction of H3N2 into the population (Fig.3). The highest average attack rate was observed in the pandemic year (mean 47.7% [CrI 47.5%, 48.0%]), the year that the virus was introduced into the human population in which there was (assumed to be) no immunity. Following the pandemic year there was a reduction in attack rate into the second (28.6% [27.8%, 29.4%]) and then third epidemic year (16.9% [15.7%,18.2%]) in which it reached a minimum. Following this minimum, the average annual attack rate gradually increased, reaching an equilibrium approximately 80 years after the pandemic year (Fig.3A). At 80–89 years since the pandemic the average annual attack rate was 22.1%, [21.5%, 22.7%], almost 23% greater than the average annual attack rate of 18.0% [17.5%, 18.5%] seen 2–9 years after the pandemic (Fig.3B). This relative increase in the average attack rate over time is sensitive to the assumed average contact rate (between individuals) in the transmission model with higher (lower) contact rates leading to greater (lesser) relative increases in the attack rate (Sup Fig.6 and Sup Fig.7).

**Figure 3.**
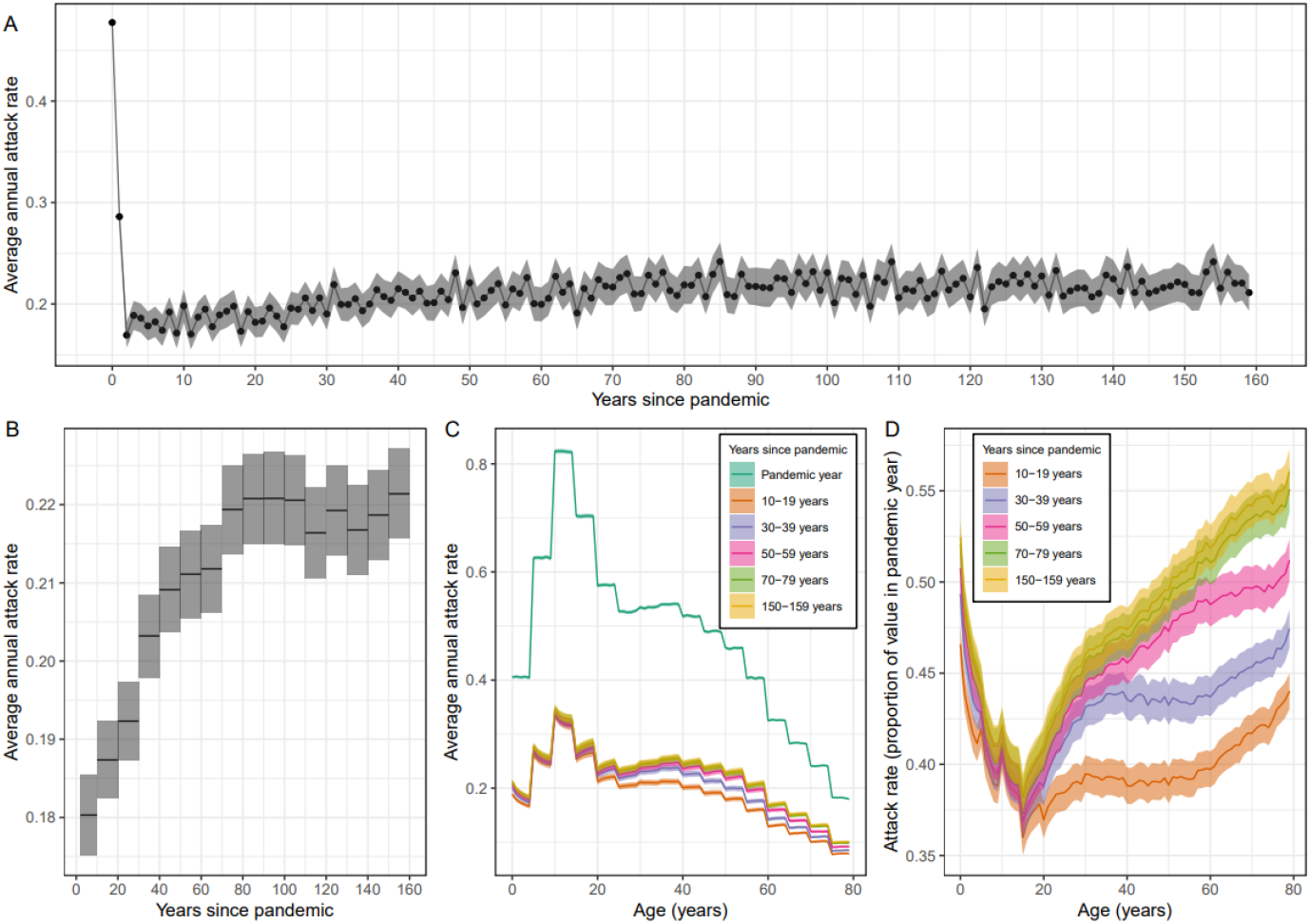
Long-term changes in the average annual attack rate of influenza. **(A)** Average annual attack rate for each epidemic year, including the pandemic year (years since pandemic = 0). Estimates are shown for the mean (points) annual attack rate and the 95% confidence interval of the mean (shaded region). **(B)** The mean annual attack rate (black line) and 95% confidence interval in the mean (shaded region) calculated instead for each decade. The first ‘decade’ only includes the years 2–9 years after the pandemic (i.e., does not include the first two years in which the attack rate had not yet reached its minimum). **(C)** The average annual attack rate by age (0–79 years) for the pandemic year, and for specific decades. Estimates are again shown for the mean (central line) and the 95% confidence interval in the mean (shaded regions). **(D)** The average annual attack rate by age for the same decades as in (C) but now shown as a proportion of the mean annual attack rate in the pandemic year.

There was significant variation in the average annual attack rates by age (Fig.3). Large differences were observed between age-groups (five-year bands), reflecting differences between their assumed contact rates. In the pandemic year, there was little within-age-group variation in the average attack rate (Fig.3C). This is a result of all individuals initially having the same immune profile (assumed to be no immunity). In later years there was within age-group variation. For those aged 0–15 years, there were lower attack rates in older individuals (within an age-group). In those aged over 15 years, there were higher attack rates in older individuals (within an age-group). The pattern in those aged 0–15 years changed little over time (Fig.3D), likely reflecting the relatively quick demographic process causing it; younger individuals have less immunity and so relatively greater attack rates. In contrast, the increase in attack rates for older individuals becomes more pronounced over time, reflecting a slower demographic process; antigenic seniority results in a reduced immune response with each successive infection over time. As time since the pandemic year increases the relative attack rate in older individuals increases to a greater extent, only reaching equilibrium after approximately 80 years. Again this relative increase in attack rates was sensitive to the assumed average contact rate (Sup Fig.6 and Sup Fig.7).

### Cohort effects

We identified differences in the number of infections over an individual’s lifetime based on their year of birth (Fig.4). Of course, those born before the pandemic year have a smaller number of infections over their lifetime because influenza circulated for a lower proportion of their lifetime. Therefore, we only considered cohorts (by year of birth) who experienced an entire lifetime of influenza circulation. Individuals born two years after the pandemic had the lowest expected number of lifetime infections, 16.6 [16.4, 16.9], which was only slightly lower than for individuals born in the pandemic year who had 16.9 [16.7, 17.1] infections over their lifetime. Considering only cohorts born three or more years after the pandemic, the expected number of lifetime infections increased slightly with year of birth reaching 17.6 [17.4, 17.7] for those born 80 years after the pandemic, an increase of approximately 6% from the minimum value.

**Figure 4.**
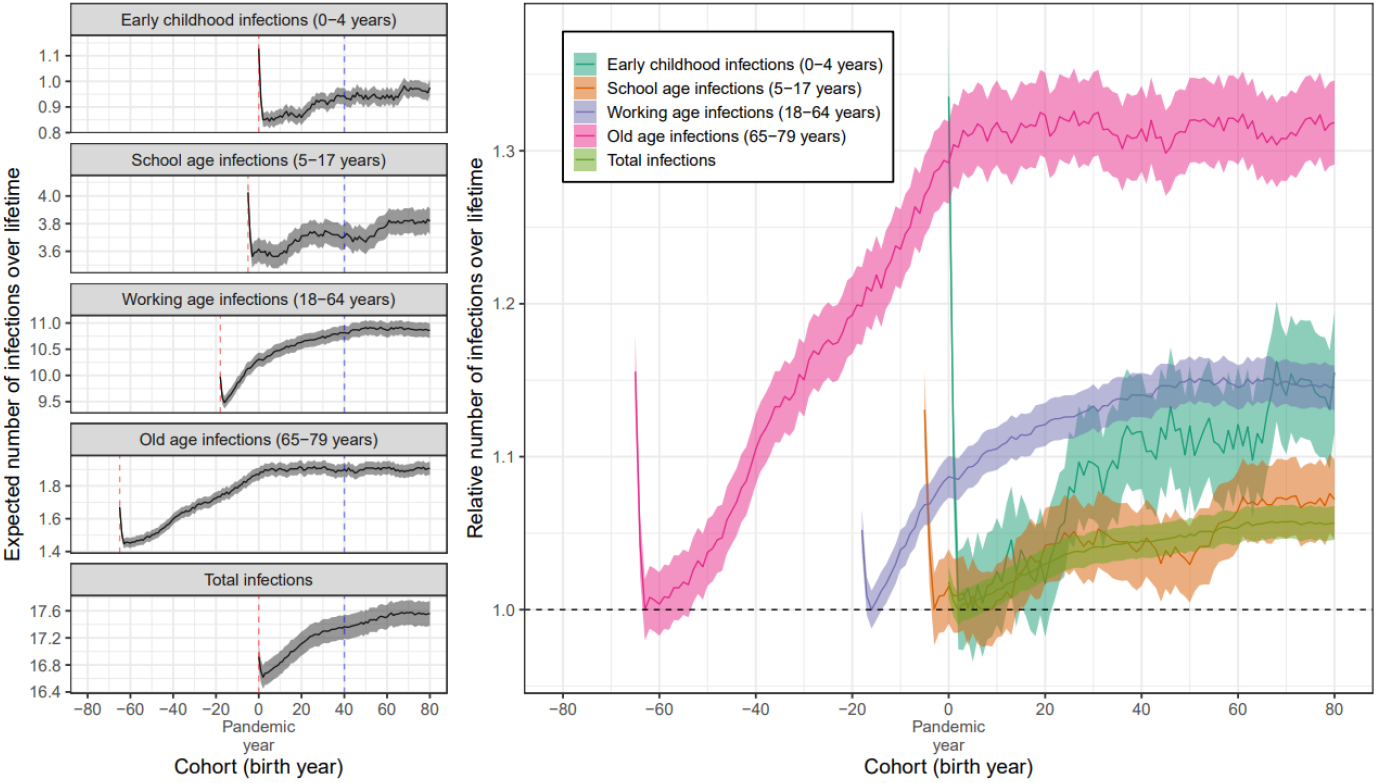
The expected number of infections by year of birth. (Left) The expected number of infections an individual will experience during different life-stages (early childhood, school age, working age, and old age) and overall (total) by their year of birth (cohort). For example, an individual born 40 years after the pandemic (blue dashed line) will on average experience 0.94 early childhood infections, 3.71 school age infections, 10.81 working age infections, and 1.90 old age infections, for a total of 17.37 infections over their lifetime. Estimates are only shown for cohorts where influenza was circulating all years in which they were in the life-stage (e.g., for total infections only those born in the pandemic year or later, and for old age infections those born 65 years before the pandemic or later). The red dashed line shows the cohort which entered the life-stage in the year of the pandemic. (Right) The expected number of infections an individual will experience during different life-stages (and overall) by cohort relative to the cohorts for which the value (for each life-stage) is a minimum (black dashed line). All estimates are shown with the mean (lines) and the 95% confidence interval in the mean (shaded region).

There were substantial differences in the number of infections an individual was expected to experience during any specific life stage depending on their year of birth. Based on broad mixing patterns and the potential consequences of infection, we considered four life stages: early age (0-4 years old), school age (5-17 years old), working age (18-64 years old) and old age (65-79 years old). For all life stages considered, the cohort entering the life stage in the year of the pandemic had an increased expected number of infections during that life stage. Similarly, the cohort entering the life stage two years after the pandemic had the lowest expected number of infections during that life stage. These results would be the same for other specified life stages (Sup Fig. 8), reflecting the increased attack rates observed in the pandemic year (and the year after the pandemic), and the relative minimum in attack rates seen two years after the pandemic (i.e., the cohort entering a life stage two years after the pandemic experiences this minimum in attack rates).

The number of infections an individual was expected to experience during any specific life stage increased with their year of birth. We only considered the cohorts entering a life-stage two years after the pandemic (when the relative minimum in expected number of infections occurred) or later. There were modest increases in the number of expected school age (5–17 years) infections with the cohort born 80 years after the pandemic (last year considered) expecting 3.82 [3.73, 3.91] infections compared to the minimum of 3.56 [3.48, 3.65] infections (cohort born two years before the pandemic). The expected number of working age (18–64 years) and early-age (0–4 years) infections both increased by approximately 15% from their minimum values to 10.9 [10.7, 11.0] and 0.98 [0.94, 1.01] infections respectively for the cohort born 80 years after the pandemic. Finally, the expected number of old-age (65–79 years) infections increased greatly with those born in the year of the pandemic (after which an approximate equilibrium was reached) or later expecting over 30% more old-age infections than the minimum (for those born 63 years before the pandemic). For those born 80 years after the pandemic the expected number of old age infections was 1.91 [1.87, 1.95]. Direct comparisons between the specific life stages we considered are hampered due to their different sized age-ranges, but when considering the cohort dynamics for life stages with equally sized age-ranges (five-year age bands), greater relative increases in the expected number of infections were observed for older-aged infections (Sup Fig.8).

### The role of antigenic seniority, short-term immunity and long-term immunity

The long-term dynamics of influenza were highly dependent on the assumed degree of antigenic seniority. When no antigenic seniority was assumed, the average annual attack rate reached an equilibrium just a few years after the pandemic, and did not increase thereafter (Fig.5A). Conversely, a higher degree of antigenic seniority resulted in a greater total increase in the average annual attack rate and a longer time to reach an equilibrium.

**Figure 5.**
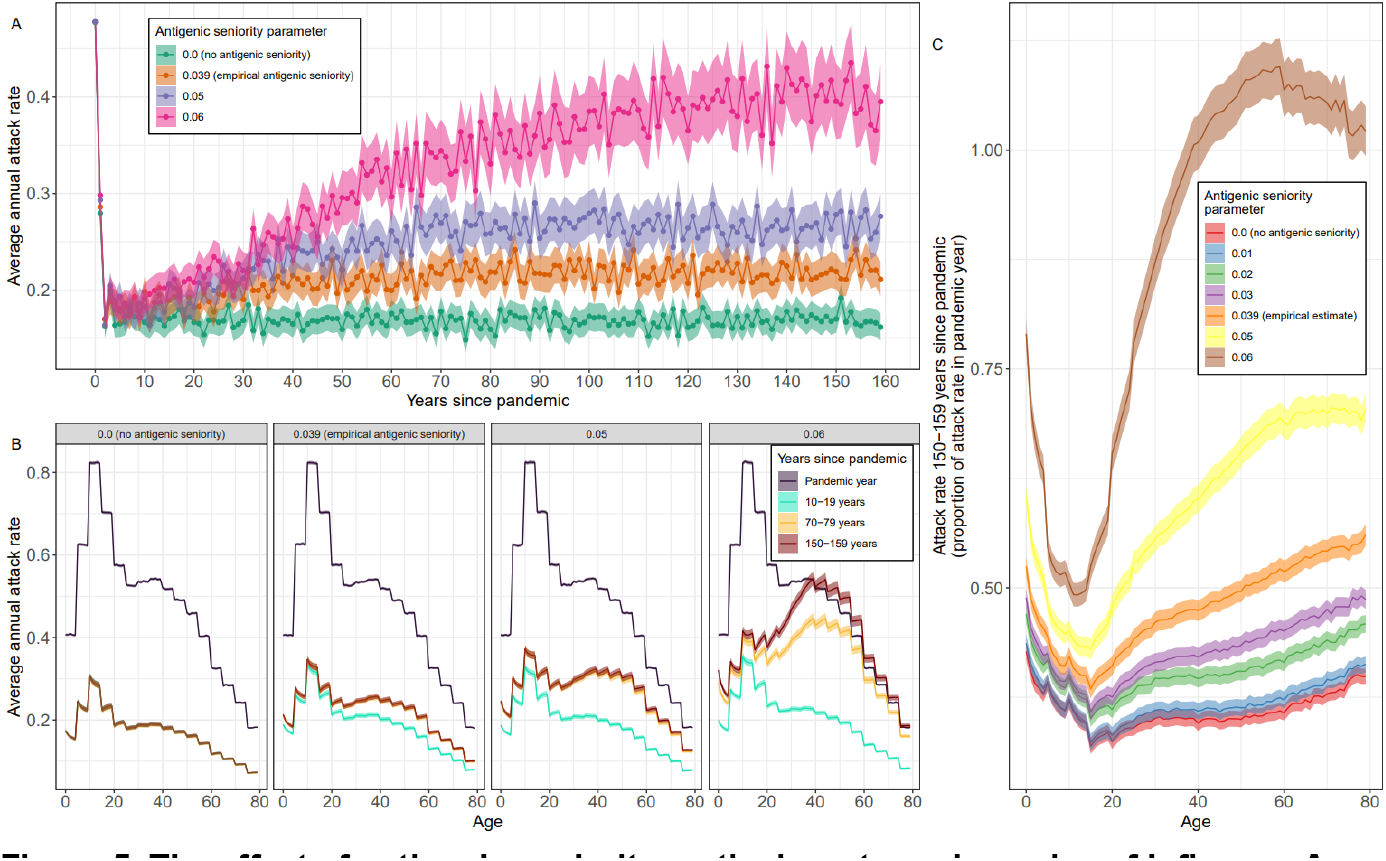
The effect of antigenic seniority on the long-term dynamics of influenza A H3N2. **(A)** The average annual attack rate for each epidemic year, including the pandemic year (years since pandemic = 0), for different values of the antigenic seniority parameter (see Methods). The number of infections until the antibody response is suppressed by 50% is given by 0.5 divided by the antigenic seniority parameter. Estimates are shown with the mean (points) and the 95% confidence interval for the mean (shaded region). **(B)** The average annual attack rate by age (0–79 years) for the pandemic year, and for specific decades, for different values of the antigenic seniority parameter. Estimates are shown with the mean (central line) and the 95% confidence interval for the mean (shaded regions). **(C)** The average annual attack rate by age for the years 150–159 years after the pandemic, shown relative to the average annual attack rate by age in the year of the pandemic, for different values of the antigenic seniority parameter. Estimates are again shown with the mean (central line) and the 95% confidence interval for the mean (shaded regions).

Antigenic seniority had little effect on the dynamics over the first ten years, with similar average attack rates for different values of antigenic seniority. However, for higher degrees of antigenic seniority there were greater increases in the average annual attack rate and after 150 years there were substantial differences. With no antigenic seniority the attack rate 150-159 years after the pandemic (Sup Tab.1) was 17.0% [16.5%, 17.4%]. With antigenic seniority representing a 50% reduction in antibody response after 13, 10 and 9 infections the attack rate 150–159 years after the pandemic was 22.1% [21.6%, 22.7%], 26.9% [26.1%, 27.6%] and 39.5% [38.3%, 40.7%] respectively. The increase in attack rate was again proportionally higher in older individuals (Fig.5C, Sup Fig.9).

The average annual attack rate was highly dependent on short- and long-term antibody responses. Increasing the strength of long-term immunity, or the duration of short-term immunity, led to lower levels of the average annual attack rate (Fig.6 and Sup Fig.10). This in turn had a feedback effect on the long-term dynamics; when the average attack rate is higher individuals are infected more often over their lifetime and so antigenic seniority has a more pronounced effect, driving greater increases in the attack rate over time.

**Figure 6.**
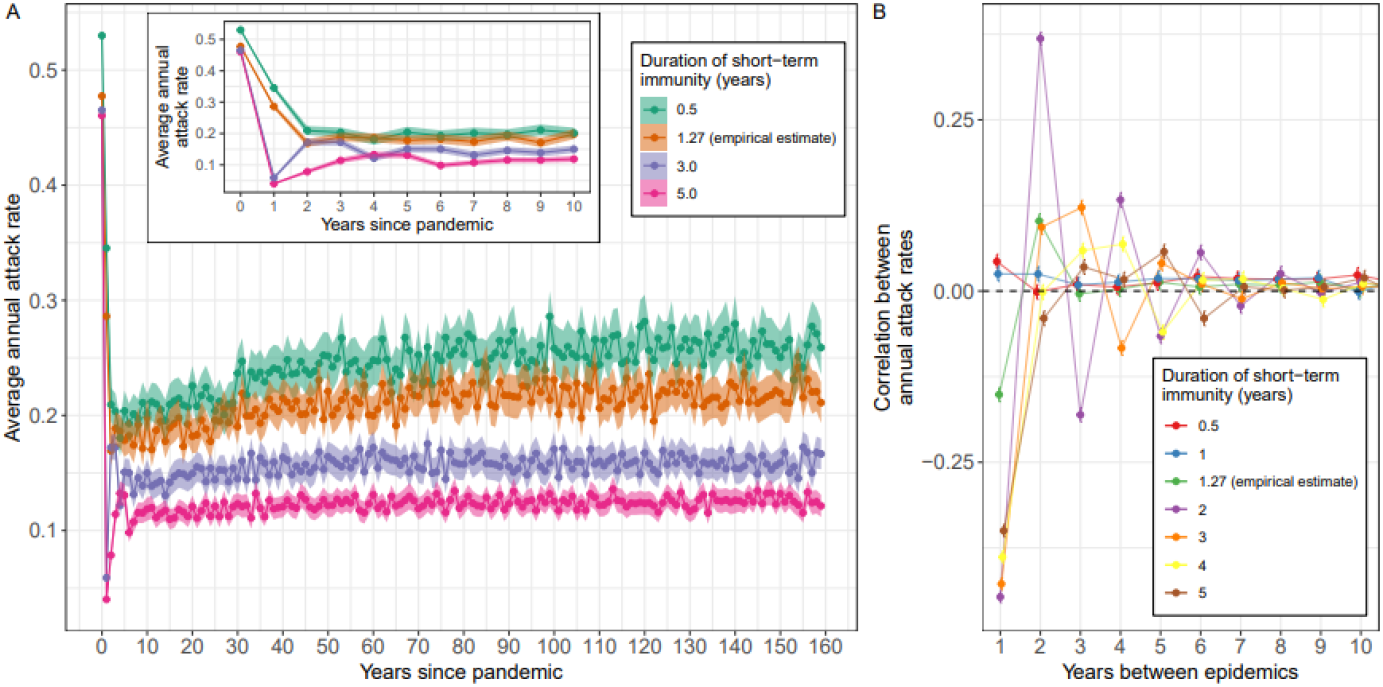
The role of short-term immunity in the epidemiological dynamics of influenza A H3N2. **(A)** The average annual attack rate for each epidemic year, including the pandemic year (years since pandemic = 0), for different durations of short-term immunity (time before short-term immunity goes to zero). Estimates are shown with mean (points) and central 95% confidence intervals (shaded region). **(B)** The average correlation in the annual attack rate between epidemic years (as a function of the number of years between epidemics), estimated over the epidemic years 20–159 years after the pandemic. Estimates are shown with their mean (points) and 95% confidence interval (bars) for different durations of short-term immunity (colours, see legend). The dashed black line highlights a correlation of 0 (no correlation).

Comparing across simulations we were able to identify an autocorrelation structure (Sup Fig.5). We measured the correlation of annual attack rates between epidemic years. For the empirical estimate of all immunity parameters, there was a significant level of negative correlation, -0.15 [-0.16, -0.14], between adjacent years; large epidemics were more likely to be followed by small epidemics. In contrast, epidemics separated by two years had a significant level of positive correlation, 0.10 [0.09, 0.11]; large epidemics were more likely to be followed by large epidemics two years later (probably due to small epidemics being more likely in the year between). There was little difference between the correlations for epidemics separated by 3–20 years, with a small level of positive correlation of approximately 0.01. This auto-correlation structure was approximately stable from 20 years after the pandemic (Sup Fig.5).

Changing the duration of short-term immunity influenced the auto-correlation structure between epidemics (Fig.6B). There was a greater level of negative correlation between adjacent epidemics when the duration of short-term immunity was longer. The relationship was more complex for epidemics separated by multiple years; for example, for epidemics separated by four years increasing the duration of short-term immunity to two years (the empirical estimate is 1.27 years) led to positive correlation but increasing the duration of short-term immunity to 3 years led to negative correlation. The changes in autocorrelation structures were only apparent in the average annual attack rates during the first ten years of the simulation (Fig.6A).

## Discussion

By integrating models of transmission, antigenic drift, and immunity, we have simulated the long-term dynamics of influenza H3N2 in temperate regions. While previous studies have focused on the evolutionary dynamics of influenza [11–13], we have explored the long-term epidemiological dynamics of H3N2 transmission. Our analysis considers influenza dynamics over a 160-year timescale, reflecting circulation of H3N2 since its emergence in 1968 during the Hong Kong pandemic through to the year 2128. Our model captures the stochastic nature of influenza transmission with high annual variability in the epidemic attack rate. We observed that on average the annual attack rate should have been increasing since 1970 (two years after the pandemic), with relatively greater increases in the annual attack rate for older individuals. This is difficult to verify empirically, as data on influenza infections have not been collected consistently since H3N2’s emergence. And what data are available are often heavily biased towards individuals seeking healthcare (who are not representative of the overall population of infected individuals) [16].

Our findings have implications for public health. If the average annual attack rate for H3N2 has still not reached an equilibrium, as suggested by our analysis (where the equilibrium is expected only after 80 years, i.e., around the year 2048), then increases in the average annual attack rate may be expected in the coming decades. This could drive an increase in the future healthcare demand due to H3N2 infections, which would be compounded by relatively greater increases in infections for older individuals, for whom the risk of severe illness is highest [17]. Additionally, our analysis highlights differences between individual cohorts: someone born in 1968 (or later) was expected to have over 30% more infections in their old age (65–79 years) than someone who was 63 in the year of the pandemic (born in 1905). Future individual risk to severe (old age) infections has thus already reached its maximum in those currently aged under 55 years old.

The model makes several simplifying assumptions. Countries’ age-distributions vary due to demographic factors [18]. Our results, which assume a uniform age-distribution, must be considered in this context given the sharp increase in severity of infection with age [19]. Age-distributions may also change over time (with many countries having ageing populations), influencing epidemic dynamics [20]. This could potentially introduce more complex cohort effects in the context of the multi-strain transmission dynamics considered here. Similarly, all contact rates were modelled as constant over time; a long-term trend of increasing contact rates would lead to additional upwards pressure on annual influenza attack rates (and vice versa). There have been some short-term reductions in contact rates due to interventions introduced during the SARS-CoV-2 pandemic [21], these measures reduced influenza transmission for much of 2020–2022 [22]. The potential cohort effects for H3N2 epidemiology introduced by responses to COVID-19 will likely not be immediately apparent. Our model considers only infection with influenza H3N2, but there have been many respiratory viruses (e.g, influenza H1N1) circulating which can interfere with transmission [23,24]; this might have acted to reduce attack rates from what would have been observed without their circulation/interference.

We found that the long-term dynamics of influenza H3N2 were highly dependent on the degree of antigenic seniority. Higher levels of antigenic seniority resulted in a greater increase in the average annual attack rate. If there was no antigenic seniority, then the average annual attack rate would have reached an equilibrium shortly after the pandemic and would not have increased thereafter. We used a simple parameterisation of antigenic seniority using the results of Kucharski et al [9]. This is the only available parameterisation of antibody dynamics that includes antigenic seniority and relates the antibody response between two strains to their antigenic separation (necessary for our model formulation). A different parameterisation of antigenic seniority used an exponential function [14] but given the expected total number of infections of an individual in our model (approximately 17–18 over a lifetime), the two functions would not be expected to produce substantially different results (these two choices of functional forms are more divergent at greater numbers of infections). Both parameterisations are based on serology studies collecting data approximately 40 years after the pandemic, and so the validity of these models after 80 years’ of virus circulation is unclear; though this would be a problem for any recent dataset given the long timescale of our analysis.

The magnitude of effect that antigenic seniority had on the long-term dynamics of influenza H3N2 was heavily modulated by the average annual attack rate in our model. Changing the overall level of immunity conferred by infection — either by increasing the strength of long-term immunity or by reducing the waning rate of short-term immunity — led to a change in the baseline average attack rate. With higher annual attack rates, individuals had a greater number of infections over their lifetime, and so antigenic seniority had a greater impact on the long-term dynamics. The average annual attack rate of influenza H3N2 is poorly characterised. We selected model parameters so that the average annual attack rate over the first 40 years of simulations was approximately 20%. This is consistent with results from analysis performed using longitudinal serology data on a cohort in Vietnam [9], although estimates of the attack rate from another Vietnamese cohort were higher at 25.6% [25]. However, it is not clear if this is an appropriate value for the temperate regions considered in this paper. Some estimates available for temperate regions have been lower [26], but the methodology used primarily identified symptomatic cases and it is known that a high proportion of influenza infections are asymptomatic [27]. Studies using the ‘gold standard’ method for identifying recent influenza infection (serologic detection of a four-fold increase in antibody titres) have estimated greater overall annual attack rates for seasonal influenza (H3N2, H1N1, influenza B) of 19% in the United Kingdom [27] and 24% in New Zealand [28], of which H3N2 should be a significant proportion [5]. However, another study has suggested that even when these ‘gold-standard’ methods are used, they could be underestimating attack rates by as much as 27% [29].

We did not consider the effect of vaccination. Influenza vaccines are updated regularly to better match circulating strains [30], and are deployed annually in many countries with varying rates of coverage [31]. While vaccination can prevent illness, hospitalisations, and deaths (when the vaccine strain is a good match to circulating strains) [32,33], the effect on short-term transmission dynamics in the population may be minimal: subsets of the population with higher vaccination coverage (older individuals) [34] often contribute less to transmission [35]; vaccines have limited effectiveness when they match poorly to the circulating strains [32], which is a regular occurrence for H3N2 due to its rapid antigenic evolution; and though vaccines can be effective at preventing symptomatic infection, their effectiveness against asymptomatic infection is poorly understood [36]. However, vaccination may introduce long-term effects that we are not able to consider, since the effect of vaccination on an individual’s long-term immune response is not well understood. This includes the degree to which vaccination influences antigenic seniority. We note that some work has identified reduced antibody responses following vaccination, in individuals who are vaccinated frequently [37].

Although our model has been calibrated for influenza H3N2, it could be generalised to other pathogens that have similar dynamics in which antigenic drift sustains long term transmission. Influenza H1N1pndm09 has circulated in the human population since the 2009 ‘Swine flu’ pandemic and likely exhibits similar mechanisms of immunity [38] (though this has not been confirmed). During the 2009 pandemic there were more complex patterns of pre-existing immunity to H1N1pndm09 due to: 1) the circulation of other H1N1 strains from 1977–2009 (going extinct following the 2009 pandemic) [1,39]; and 2) the antigenic similarity of the H1N1pndm09 strain to the 1918 pandemic H1N1 strain [2], meaning that older individuals (who may have been infected with more antigenically similar strains) had higher levels of pre-existing immunity [40]. It is thus unclear at what stage of its long-term dynamics the H1N1pndm09 virus is currently at. Is it already at an equilibrium state due to high levels of previous exposure to H1N1 viruses? Or given it is less than 15 years since the pandemic, is the annual attack rate still at a relative minimum? The dynamics of SARS-CoV-2 are even less well understood. Since its introduction to the human population in late 2019, we have seen the frequent emergence of new immune-evading variants. A preliminary analysis using antigenic cartography has identified that these new variants are antigenically distinct to previous variants [41]. However, it is still unclear what long-term dynamics of immunity and evolution will be exhibited by SARS-CoV-2 and if they will resemble that of influenza H3N2. Recent serological studies have demonstrated dampened immune responses due to repeat exposure to SARS-CoV-2 [42,43], hinting at the presence of antigenic seniority, but this requires investigation over a longer time frame.

Our study highlights the need for data. Following a pandemic, our analysis suggests that a relative minimum in the annual attack rate may be observed, but if there is any degree of antigenic seniority governing the immune response to future infections, then the average annual attack rate will increase over demographic timescales. Studies are needed to assess the dynamics of H1N1pndm09 and SARS-CoV-2 infections and immunity so that any long-term rise in infection rates can be anticipated. Quantifying the interactions between vaccination and long-term immunity is crucial (including for H3N2) for identifying cohorts that may be at greater risk of infection in the future. A better understanding of the effect of vaccination on long-term immunity would also allow vaccination strategies to be optimised against short- and long-term public health objectives at both the population- and individual-level. Developing long-term surveillance strategies for respiratory viruses [44], that consistently estimate population infection levels, antigenic evolution and immunity, would allow the long-term dynamics to be better quantified and public health strategies to be refined.

## Methods

### Global drift model

The antigenic drift of influenza was simulated across a two-dimensional strain space, analogous to an antigenic map. Individual strains were defined by their two-dimensional antigenic coordinates,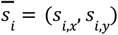. In a given year, *N*, strains were assumed to be normally distributed around a mean coordinate,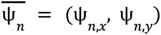, with standard deviation, ρ:

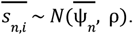

Between years the mean coordinate drifts stochastically:

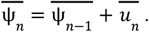

Where,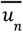, is a random variable. The x-component of 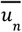 is drawn from an exponential distribution, *u* ∼*Exp*(ζ), and the y-component of 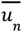 is drawn from a normal distribution, *u*_*n,y*_∼*N*(0,η). This formulation ensures that the principal component of antigenic drift is along the x-axis, while still allowing variation across both dimensions. This model of antigenic drift can be defined by the three parameters: ρ, ζ, and η.

In order to perform stochastic simulations of H3N2’s antigenic drift we first estimated the parameter values. We implemented the above model in STAN and sampled from the parameter posterior using a No-U-Turns Sampler [45]. In the implementation of the model the random variable distributions were the likelihood. All parameters were given an uninformative constant prior. The model was fitted to an antigenic map for influenza H3N2 covering the years 1968-2011 [1] (Sup Fig.1). Parameter estimates were taken as the mean value of parameter posteriors: ρ = 1.14; ζ = 0.99; η = 1.36.

### Epidemiological model

The long-term epidemiological dynamics of influenza were simulated using an agent-based multi-strain transmission model following a formulation of the SIS model [46]. At any point in time, an individual (agent) was either susceptible or infected (and infectious) (with a specific strain). Discrete epidemic years (each one 365 days long) were simulated with daily timesteps. During each timestep, either a susceptible individual could become infected following contact with an infected individual, or an infected individual could clear their infection and re-enter the susceptible class (though the individual would now have immunity to infection with antigenically similar strains, see below). Infection histories (strain and timing of infections) were tracked for all individuals.

Infected individuals cleared their infection (reentered the susceptible class) at a constant rate γ. During each timestep the number of infected individuals who cleared their infection was given by a random number chosen from a Poisson distribution, *Pois*(*I*_*t*_×γ), where *I*_*t*_ was the number of infected individuals on the given day. Infected individuals were selected to clear their infection at random. The parameter γ was chosen to have a value of 0.33 to give an average duration of infection of 3 days.

Contact between individuals was mediated by their age-groups. At the beginning of the simulation each individual was assigned an integer age with equal proportions aged 0 to 79 inclusive (1000 individuals each age for a total population of 80,000). Age-groups were defined as 5-year age bands (0–4 years, 5–9 years, etc.). Relative contact rates between age-groups were calculated using the R package conmat [47]. The relative contact rates were scaled so that the maximum contact rate was equal to 1 (the mean contact rate was 0.0742) (Sup Tab.2). Contacts between infectious individuals and susceptible individuals were then calculated using a similar approach to Tsai et al [48]. The maximum number of contacts was given by a random number chosen from a Poisson distribution,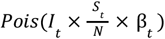 Where *I*_*t*_ is the number of infectious individuals at time,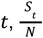 is the proportion of the population susceptible and β_*t*_ is the seasonally forced maximum contact rate. β_*t*_ is assumed to be sinusoidally varying with a period of one year, an average value of β_0_ and a maximum value of β_0_ + β_1_ : β_*t*_ = β_0_×(1 − β_1_ cos(2π*t*/365)). Given a maximum number of contacts, pairs of infectious and susceptible individuals are randomly selected. The probability of an infection event in a pair is then given by:

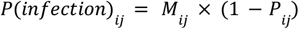

Here the probability that susceptible individual *j* is infected by infectious individual *i* is controlled by *M*_*ij*_, the contact rate between *i* and *j*’s age-groups, and *P*_*ij*_, the susceptibility of individual *j* to the strain infecting individual *i* (see Immunity model below).

Strains are introduced into the epidemic model from the global pool model. Each daily timestep a random number of susceptible individuals are selected using a Poisson distribution, *Pois*(*S*_*t*_×χ), where χ is the importation rate parameter and was chosen to be small (χ = 0. 00005). Each individual selected is then infected with a strain (randomly selected from the global pool) with probability 1 − *P*_*ij*_ . Where *P*_*ij*_ is again individual *j*’s protection against infection with strain *i*.

Between epidemic years: (1) the global drift model is updated (see above), (2) each individual’s age increases by 1 and those that should have turned 80 are reset to 0 year olds with no infection history (birth and death), and (3) all current infections are cleared. Clearing infections is done so that strains from previous years are removed when the global drift model is updated. At this point in the epidemic year (end/beginning) seasonality was at its lowest and so infection levels are low.

### Immunity model

An individual’s protection against infection with a specific strain is determined by their entire infection history. An individual’s antibody concentration (measured by log-titre) against a specific strain is determined using a model presented in Kucharksi et al [9]. Each infection in an individual’s infection history produces an antibody response. The log-titre of the antibody response is controlled by multiple components:

1. Long-term boosting. Infection with a strain *i* will produce a fixed log-titre against that strain, with parameter value μ_1_.
2. Short-term boosting. Infection with a strain *m* will produce an additional contribution to the log-titre against the strain that wanes over time. The contribution is given by μ_2_ *w*(*m*) = μ_2_×max {0, 1 − ω*t*_*m*_ }, where μ_2_ is the boosting parameter, ω is the waning parameter, and *t*_*m*_ is how much time has elapsed since infection with strain *m*.
3. Cross-reactivity. Both long- and short-term boosting will also produce log-titres against antigenically similar strains. The log-titre against an infecting strain *j*, due to an infection with strain *m* is given by *d*(*m, j*) = max {0, 1 − σ×δ_*mj*_ }, where δ_*mj*_ is the euclidean distance between strains *m* and *j* in two-dimensional antigenic space, and σ is a parameter that has different values for the cross-reactivity due to long-term boosting (σ_1_) and due to short-term boosting (σ_2_).
4. Antigenic seniority. An individual’s initial infection produces the response described above. For subsequent infections the antibody response is progressively suppressed. The log-titre produced by infection with strain *m* is scaled by the factor *s*(*X, m*) = max {0, 1 − τ(*N*_*m*_ − 1)}, where *N*_*m*_ is the number of strain *m* in the infection history (e.g, the first strain is 1, the second is 2), τ is a further parameter of the model and *X* is the set of all strains in an individual’s infection history.

The overall log-titre against strain *j*, due to an individual’s entire infection history is then given by:

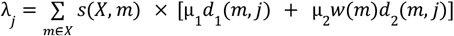

This is controlled by the parameters: μ_1_, σ_1_, μ_2_, σ_2_, ω, and τ. We take the values of these parameters from Kucharski et al. [9]: μ_1_ = 2. 02; σ_1_ = 0. 130; μ_2_ = _2_. 69; σ_2_ = 0. 031; ω = 0. 79; τ = 0. 039.

We convert the log-titre to protection against infection using the model presented in Coudeville et al. [8]. The protection against infection is given by:

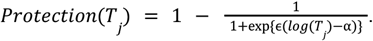

Here ϵ and α are parameters, and *T*_*j*_ is the titre against strain *j*. The conversion between *T*_*j*_ and λ_*j*_ is slightly non-trivial due to the different definitions used in the two studies considered and is given by: 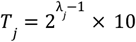 The parameter values are taken from Coudeville et al. [8]: ϵ = 1. _2_99; α = _2_. 844.

### Parameter selection

Before the long-term epidemiological dynamics of influenza transmission could be simulated, parameter values for the epidemiological model had to be selected. The parameter values have all been stated in the previous sections (and Sup Table.3) with the exception of the average contact rate, β_0_, and the degree of seasonal forcing, β_1_ (maximum contact rate of β_0_ + β_1_). We investigated the transmission dynamics for different values of these parameters. We performed 20 simulations of the first 40 years of influenza epidemics for each parameter set. For each parameter set, we measured the average annual attack rate over all simulations and years, and the variation in timing of the week in which influenza incidence was highest (peak week). We then used a generalised additive model implemented in the R package mgcv [49] to predict the value of these quantities across the continuous parameter space (Sup Fig.11). We selected parameter values so that the average annual attack rate was approximately 20% [9] and that the standard deviation in the peak week in years 21 to 40 was approximately 3.5 weeks (matching the average standard deviation in peak week timing seen in 48 US states from 1991 to 2003) [50]. The selected parameter values for simulating the long-term dynamics of influenza were β_0_ = 5. 5 and β_1_ = 0. 25. As a sensitivity analysis we also ran simulations with β_0_ = 5 and β_0_ = 5. 75; these parameters were chosen to give an average annual attack rate (in the first 40 years) of approximately 15% and 22.5% respectively.

## Statistical analyses

We simulated 160 years of influenza transmission. The main analyses were performed with the parameters given in Supplementary Table 2. Sensitivity analyses were also performed assuming different values for the antigenic seniority parameter (τ), the duration of short-term immunity (1/ω), the strength of long-term immunity (μ_1_), the average contact rate (β_0_) and for a uniform contact matrix (all contact rates between age-groups the same, equal to the mean contact rate of the contact matrix used in the main analysis) (Sup Tab.3). For each set of parameters, we ran 256 simulations (each lasting 160 years).

For each epidemic year (in each simulation) we measured the annual attack rate as the total number of infections in the year divided by the total size of the population (80,000). Similarly, we measured the annual attack rate for each integer age as the number of infections in individuals of that age divided by the number of individuals of that age (1000). We calculated the mean annual attack rate (for each year, decade and by age for specific years and decades) across simulations. The standard deviation in this mean was estimated by measuring the standard deviation in the distribution (across simulations and years [for decades]) and dividing by the square root of the sample size. The 95% confidence interval for the mean annual attack rate was estimated by subtracting/adding 1.96 times the standard deviation in the mean.

For each epidemic year we measured the Pearson correlation between the annual attack rate (in the given year) and the annual attack rate 1 to 20 years later across the 256 simulations. The standard deviation is calculated for the Fisher’s transformation of the correlation coefficient. The 95% confidence interval is then estimated by subtracting/adding 1.96 times the standard deviation and then performing the inverse Fisher’s transformation. In the first twenty years of epidemics the correlation between years changed slightly over time before reaching an equilibrium. We calculated the average correlation over all epidemic years 20 years after the pandemic year; all correlation coefficients underwent Fisher’s transformation, the mean and the standard deviation in the mean was calculated, the mean and 95% confidence intervals (+/-1.96 times the standard deviation) then underwent the inverse Fisher’s transformation.

## Data Availability

There is no data as this is a simulation study.

## Supplementary figures

**Supplementary Figure 1.**
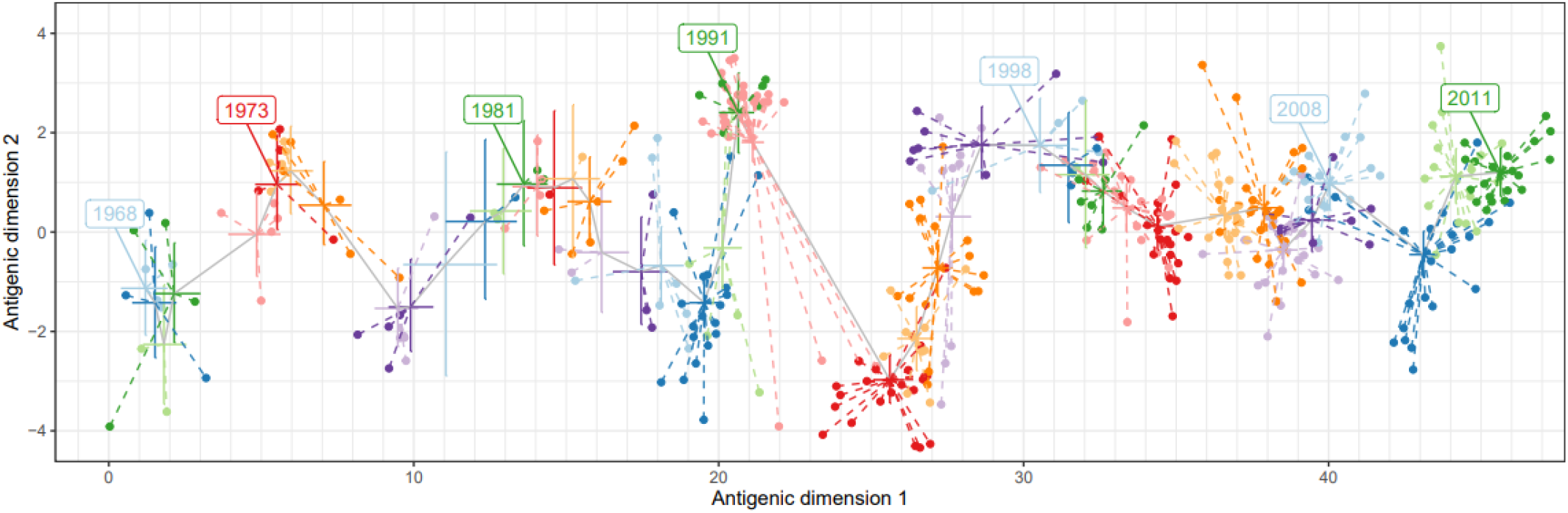
Fit of the global drift model to data from an antigenic map. Antigenic map for influenza H3N2 covering the years 1968–2011, taken from Bedford et al 2014 [1]. The antigenic distance between two strains (points) quantifies their dissimilarity, with one unit of antigenic distance corresponding to a two-fold dilution in hemagglutinin inhibition titre. Strains (points) are coloured by the year in which they were sequenced, and some years have been explicitly labelled for clarity. The statistical model describing global antigenic drift (see Methods) has been fitted to this antigenic map. The posterior of the mean antigenic coordinates,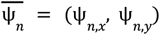 for each year are shown with the posterior mean (centre of cross) and 95% confidence interval in antigenic dimension 1 (horizontal line of cross) and antigenic dimension 2 (vertical line of cross). Grey lines connect the mean antigenic coordinates between adjacent years; note that the mean antigenic coordinates increase along antigenic dimension 1 each year (from left to right). The strains associated with a given year are connected to the mean antigenic coordinates for that year by dashed lines, and are coloured the same. Note that in some years there are no strains on the antigenic map (e.g., 1978 [the second light blue year]).

**Supplementary Figure 2.**
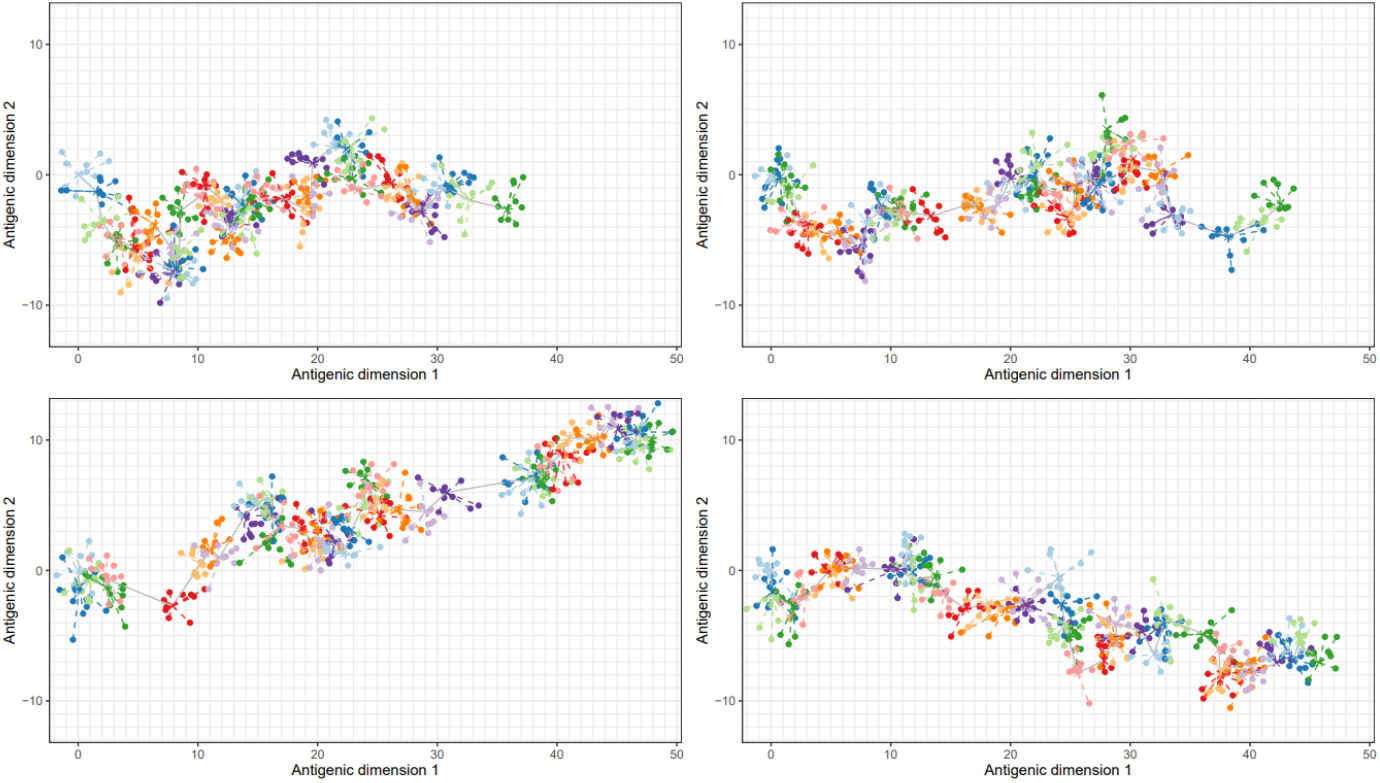
Example simulations of the global drift model. Four independent simulations of the global drift model. The simulated antigenic diversity of influenza strains for 44 years (colours highlight separate years with 10 strains per year). Strains are located on a two-dimensional antigenic map. The antigenic distance between two strains quantifies their dissimilarity, with one unit of antigenic distance corresponding to a two-fold dilution in hemagglutinin inhibition titre. Strains in a single year (points) are normally distributed around mean antigenic coordinates (dotted lines connect strains in a given year to the mean antigenic coordinates). The mean antigenic coordinates change each year (grey line connecting adjacent years), always progressing along antigenic dimension 1 (left to right). The mean antigenic coordinates in the first year were set to be (0, 0).

**Supplementary Figure 3.**
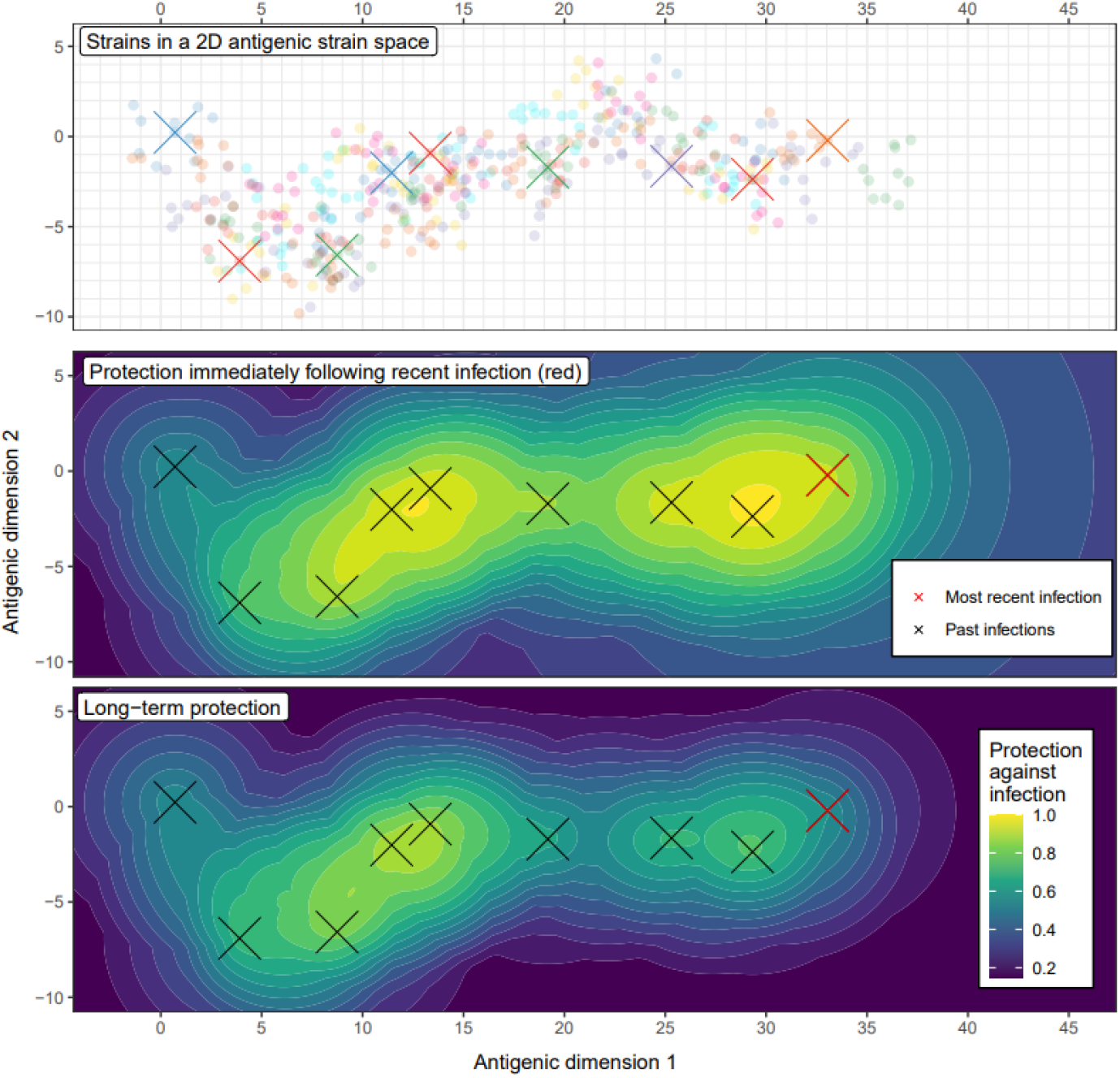
Example individual-level immunity due to multiple infections with antigenically distinct strains. (Top panel) One example simulation (see Supplementary Figure 2) of the antigenic diversity of influenza strains (points) for 44 years (colours highlight separate years with 10 strains per year). Strains are located on a two-dimensional antigenic map. The antigenic distance between two strains quantifies their dissimilarity, with one unit of antigenic distance corresponding to a two-fold dilution in hemagglutinin inhibition titre. The simulated infections of a single individual are indicated by crosses. The infection furthest along antigenic dimension 1 is the most recent infection. (Middle and bottom panels) The estimated protection against infection from a strain of influenza, as a function of the strain’s location on the antigenic map. The middle panel shows the protection against infection immediately following the most recent infection (red cross), a combination of the long-term immune responses of all past infections (black crosses and red cross), and the short-term immune response of the most recent infection. The bottom panel shows the protection against infection a long time after the most recent infection (any time greater than 1.27 years: the time taken for the short-term immune response to go to 0), a combination of only the long-term immune responses of all past infections.

**Supplementary Figure 4.**
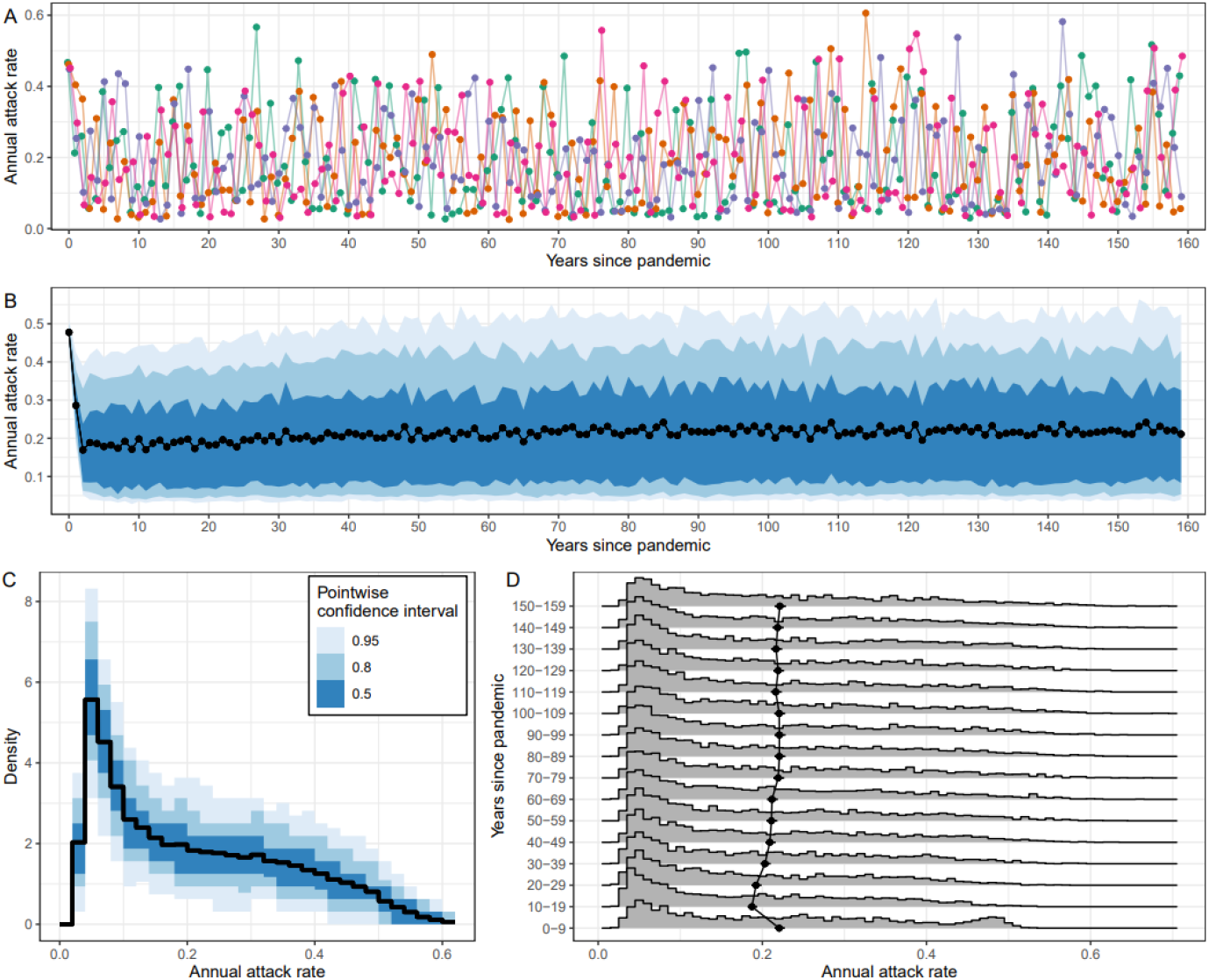
Variation in the annual attack rate between simulations. **(A)** The annual attack rate for each epidemic year (points), including the pandemic year (years since pandemic = 0), shown for four independent simulations (four unique colours and lines). **(B)** The average (mean) annual attack rate for each epidemic year (points) and the central 50% (dark blue), 80% (blue), and 95% (light blue) tolerance intervals, estimated from 256 simulations. Note that the black line and points are the same as in Figure 3. **(C)** Probability density of the annual attack rate over all 160 epidemic years of each simulation. The probability density was calculated for all 256 simulations; the median (black line) and 50%, 80%, and 95% pointwise confidence intervals (shaded regions) are shown. **(D)** Probability distributions (grey shaded regions) of the annual attack rate over all 256 simulations for each decade. Also shown are the average (mean) annual attack rates (points) and their 95% confidence interval (small black bars) for each decade.

**Supplementary Figure 5.**
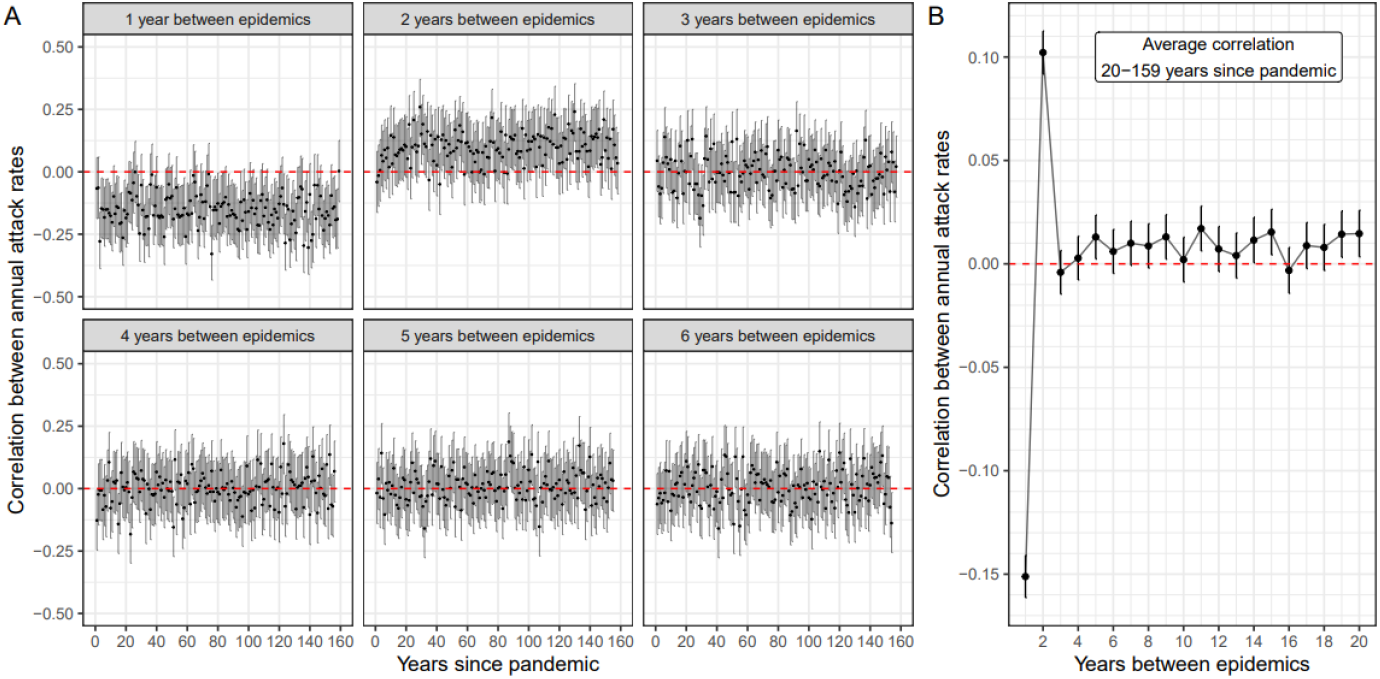
The correlation structure of the attack rate between epidemic years. **(A)** The correlation (points) between the attack rate in a single epidemic year (x-axis) and the attack rate in a future epidemic year (subheadings give time between epidemic years). Each estimate of correlation (points), and its 95% confidence interval (grey lines) is made across 256 simulations. **(B)** The average correlation of the attack rates between epidemic years (as a function of the number of years between epidemics), estimated over the epidemic years 20–159 years after the pandemic. Estimates are shown with their mean (points) and 95% confidence interval (black bars). The dashed red line in both subplots highlights a correlation of 0 (no correlation).

**Supplementary Figure 6.**
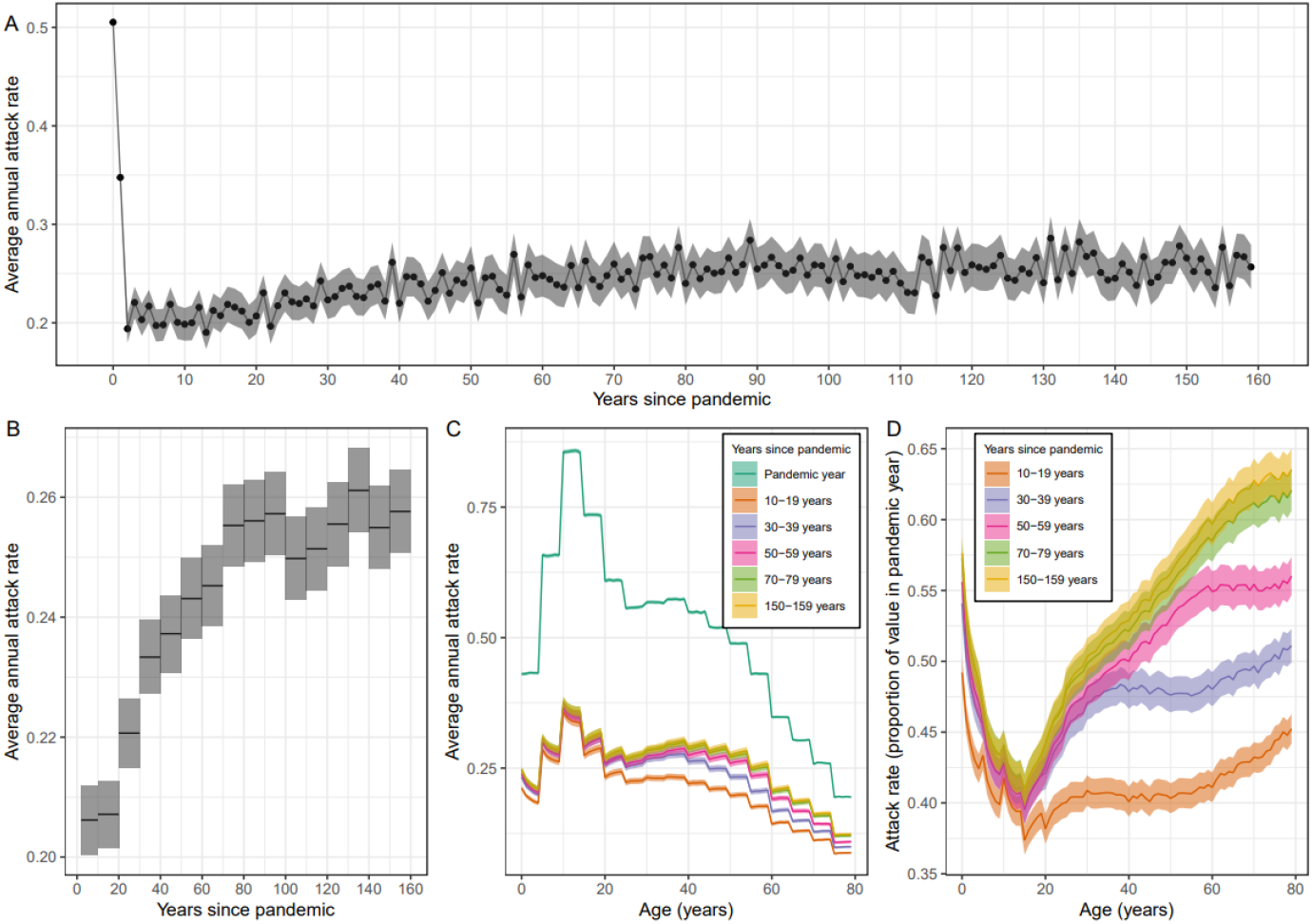
Long-term changes in the average annual attack rate of influenza. Sensitivity analysis with higher transmission rate. The same as in Figure 3, but simulations were instead run with parameters chosen so the average annual attack rate in the first 40 years was 22.5% (see Methods). **(A)** Average annual attack rate for each epidemic year, including the pandemic year (years since pandemic = 0). Estimates are shown for the mean (points) annual attack rate and the 95% confidence interval in the mean (shaded region). **(B)** The mean annual attack rate (black line) and 95% confidence interval in the mean (shaded region) calculated instead for each decade. The first ‘decade’ only includes the years 2–9 years after the pandemic (i.e., does not include the first two years in which the attack rate was still at higher levels). **(C)** The average annual attack rate by age (0–79 years) for the pandemic year, and for specific decades. Estimates are again shown for the mean (central line) and the 95% confidence interval in the mean (shaded regions). **(D)** The average annual attack rate by age for the same decades as in (C) but now shown as a proportion of the mean annual attack rate in the pandemic year.

**Supplementary Figure 7.**
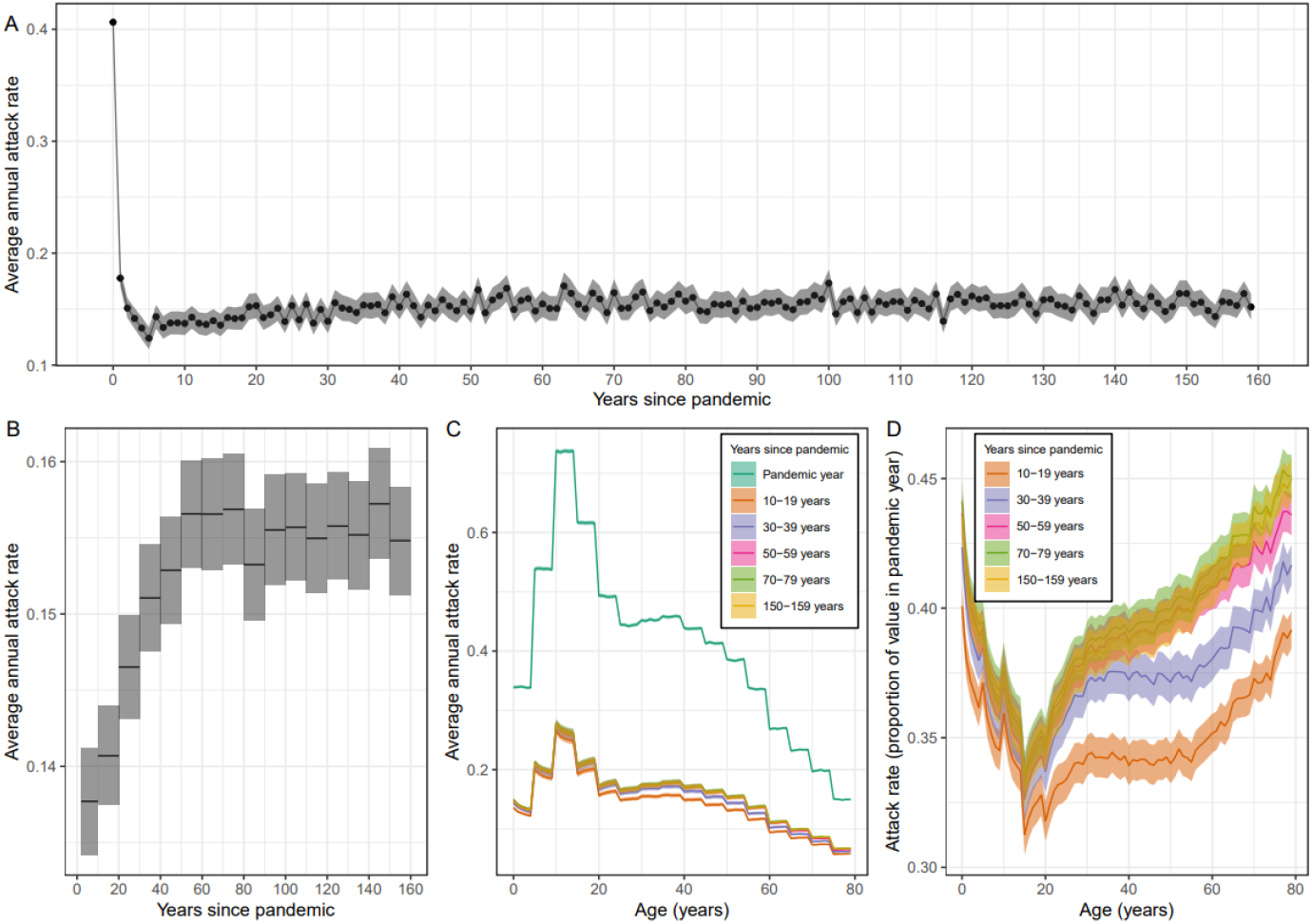
Long-term changes in the average annual attack rate of influenza. Sensitivity analysis with lower transmission rate. The same as in Figure 3, but simulations were instead run with parameters chosen so the average annual attack rate in the first 40 years was 15.0% (see Methods). **(A)** Average annual attack rate for each epidemic year, including the pandemic year (years since pandemic = 0). Estimates are shown for the mean (points) annual attack rate and the 95% confidence interval in the mean (shaded region). **(B)** The mean annual attack rate (black line) and 95% confidence interval in the mean (shaded region) calculated instead for each decade. The first ‘decade’ only includes the years 2–9 years after the pandemic (i.e., does not include the first two years in which the attack rate was still at higher levels). **(C)** The average annual attack rate by age (0–79 years) for the pandemic year, and for specific decades. Estimates are again shown for the mean (central line) and the 95% confidence interval in the mean (shaded regions). **(D)** The average annual attack rate by age for the same decades as in (C) but now shown as a proportion of the mean annual attack rate in the pandemic year.

**Supplementary Figure 8.**
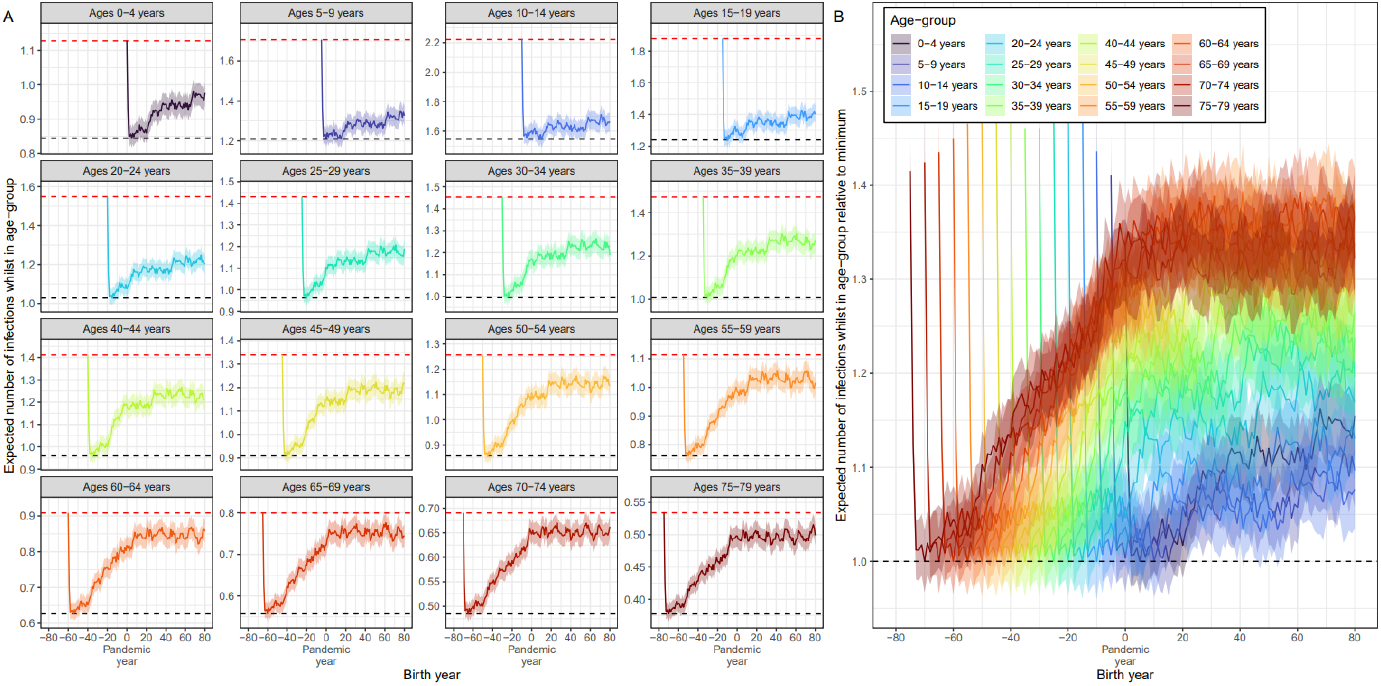
The expected number of infections by year of birth. (Left) The expected number of infections an individual will experience during different life-stages (5 year age ranges) by their year of birth (cohort). Estimates are only shown for the cohorts in which influenza was circulating all years in which they were in the life-stage. For example, for the life-stage representing infections while aged 20–24 years, only those born 20 years before the pandemic and later are considered. Horizontal dashed lines show the maximum expected number of infections, found for the cohort entering the life-stage in the year of the pandemic (red), and the minimum expected number of infections, found for the cohort entering the life-stage two years after the pandemic (black). (Right) The expected number of infections an individual will experience during different life-stages by cohort relative to the cohorts for which the value (for each life-stage) is a minimum (black dashed line). All estimates are shown with the mean (lines) and the 95% confidence interval in the mean (shaded region).

**Supplementary Figure 9.**
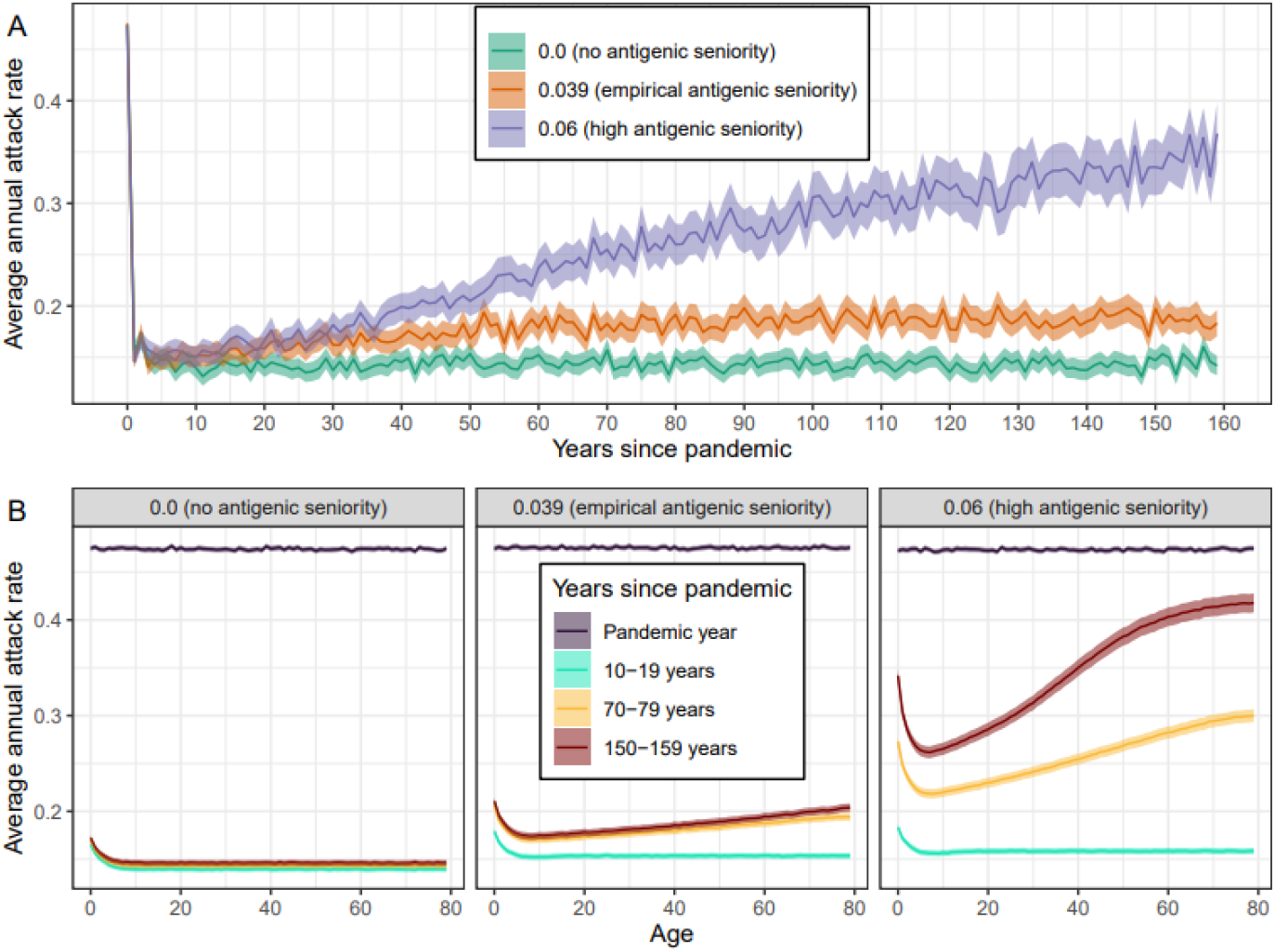
The effect of antigenic seniority on the long-term dynamics of influenza A H3N2. Sensitivity analysis using uniform contact rates. (A) and (B) are the same as in Figure 4, but the age-contact matrix now assumes equal contact rates between all age-groups (the mean contact rate matches the mean contact rate of the age-contact matrix used in the main analysis). **(A)** The average annual attack rate for each epidemic year, including the pandemic year (years since pandemic = 0), for different values of the antigenic seniority parameter (see Methods). The number of infections until the antibody response is suppressed by 50% is given by 0.5 divided by the antigenic seniority parameter. Estimates are shown with the mean (points) and the 95% confidence interval for the mean (shaded region). **(B)** The average annual attack rate by age (0–79 years) for the pandemic year, and for specific decades, for different values of the antigenic seniority parameter. Estimates are shown with the mean (central line) and the 95% confidence interval for the mean (shaded regions).

**Supplementary Figure 10.**
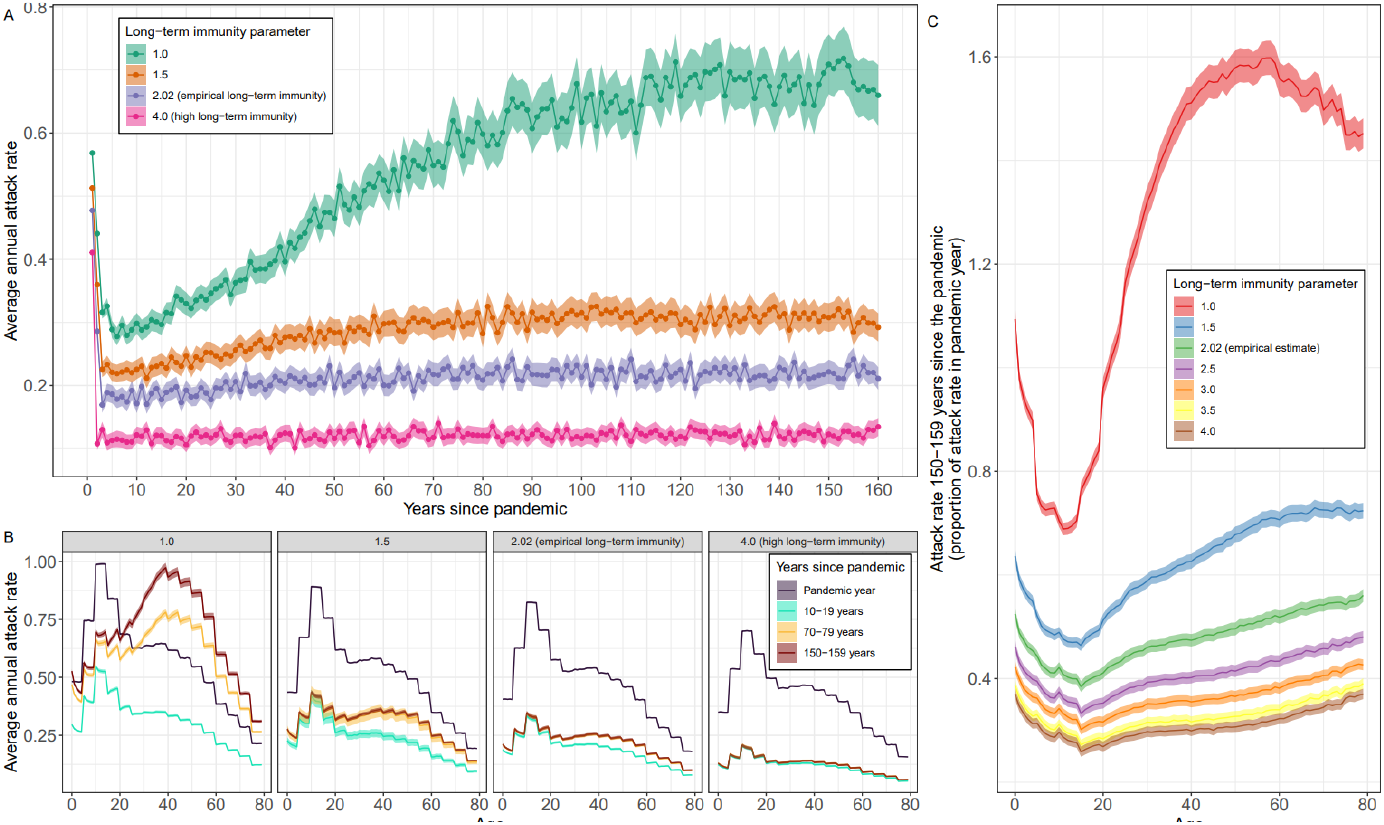
The effect of long-term immunity on the long-term dynamics of influenza A H3N2. **(A)** The average annual attack rate for each epidemic year, including the pandemic year (years since pandemic = 0), for different values of the parameter setting the maximum long-term antibody response, μ_1_(see Methods). Estimates are shown with the mean (points) and the 95% confidence interval for the mean (shaded region). **(B)** The average annual attack rate by age (0–79 years) for the pandemic year, and for specific decades, for different values of the long-term immunity parameter. Estimates are shown with the mean (central line) and the 95% confidence interval for the mean (shaded regions). **(C)** The average annual attack rate by age for the years 150–159 years after the pandemic, shown relative to the average annual attack rate by age in the year of the pandemic, for different values of the long-term immunity parameter. Estimates are again shown with the mean (central line) and the 95% confidence interval for the mean (shaded regions).

**Supplementary Figure 11.**
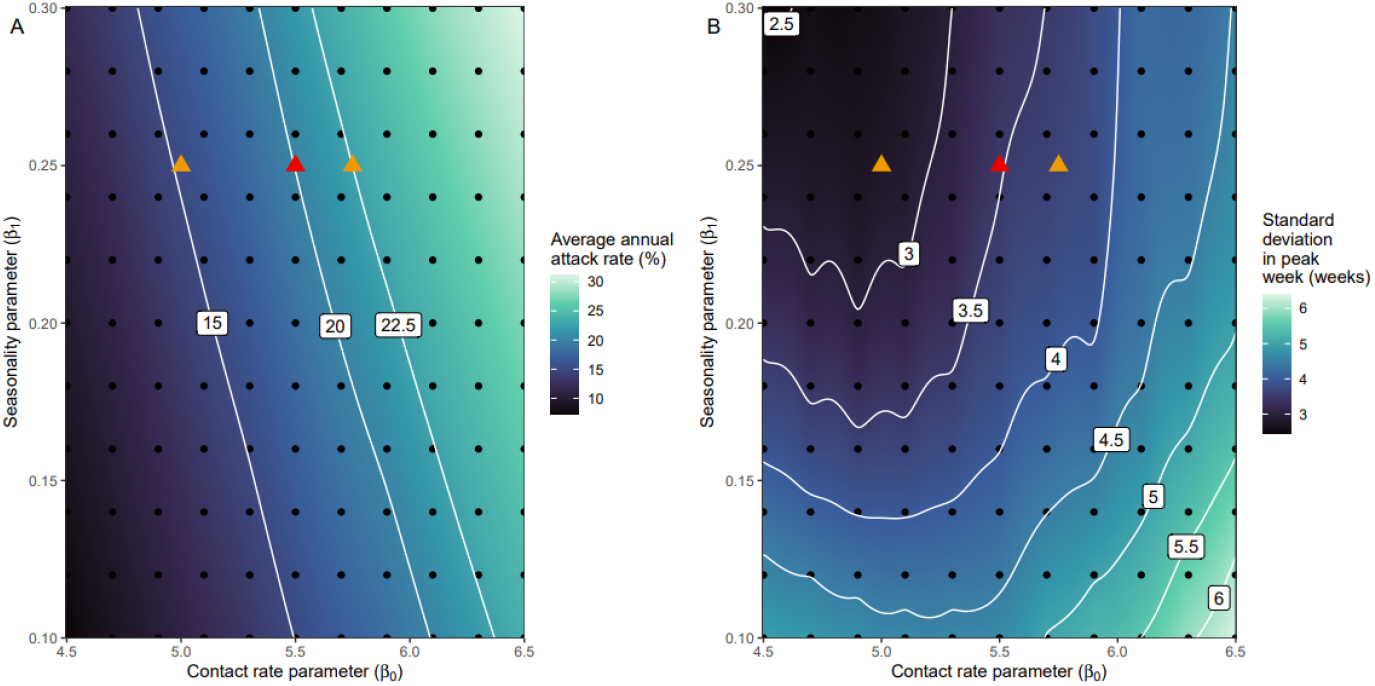
Selection of parameter values for the contact rate and seasonality. **(A)** The average annual attack rate over the first 40 epidemic years (including the pandemic year). **(B)** The standard deviation in the timing of the peak week over the first 40 epidemic years (including the pandemic year). **(A**,**B)** Point estimates were made using the epidemiological model (20 simulations) at set values (grid of black dots) of the contact rate parameter, β_0_, and the seasonality parameter, β_1_. Continuous estimates were then made using generalised additive models, with the output variable the point estimates, and the parameter values (in a two-dimensional thin plate spline) as the input variables. Contours are shown (white) for specific values of the continuous estimates. The parameters used for the main analysis (red triangle) were selected so that in the first 40 years the average annual attack rate was approximately 20% and the standard deviation in the timing of the peak week was approximately 3.5 weeks. Parameters for sensitivity analysis (orange triangles) were chosen so that in the first 40 years the average annual attack rate was approximately 15% and 22.5%.

## Supplementary tables

Supplementary tables are available in the supplementary document

**Supplementary Table 1**. Average annual attack rates for each decade, for different values of the antigenic seniority parameter.

**Supplementary Table 2**. All parameter values used in the model for the main analysis and the parameter values used (one parameter value varied at a time) in sensitivity analyses.

**Supplementary Table 3**. Contact rates between all age-groups.

